# The Impact of Management on Hospital Performance

**DOI:** 10.1101/2020.06.02.20119545

**Authors:** Miqdad Asaria, Benjamin Gershlick, Alistair McGuire, Andrew Street

## Abstract

There is a prevailing popular belief that expenditure on management by healthcare providers is wasteful, diverts resources from patient care, and distracts medical and nursing staff from getting on with their jobs. There is little existing evidence to support this narrative or counter-claims. We explore the relationship between management and public sector hospital performance using a fixed effects empirical econometric specification on a panel data set consisting of all 129 non-specialist acute National Health Service (NHS) hospitals in England for the financial years 2012/13 to 2018/19. Measures of managerial input and quality of management practice are constructed from NHS Electronic Staff Records and NHS Staff Survey data. Hospital accounts and Hospital Episode Statistics data are used to construct five measures of financial performance and of timely and high quality care. We find no evidence of association either between quantity of management and management quality nor directly between quantity of management and any of our measures of hospital performance. However, there is some evidence that higher quality management is associated with better performance. NHS managers have limited discretion in performing their managerial functions, being tightly circumscribed by official guidance, targets, and other factors outside their control. Given these constraints, our findings are unsurprising.

## 1 Introduction

Management matters, apparently. In theory, organisations that are better managed have staff that are more motivated and attuned to organisational objectives. As a consequence, these organisations ought to be more successful in pursuing their objectives than otherwise similar organisations that are less well managed. For private sector organisations evidence appears to support the theory (Bloom and Van Reenen, 2007). But when it comes to public sector organisations, governments play a significant role in determining how management is constituted, how managers are paid and what they are tasked to do. This is true for the English National Health Service (NHS), in which hospitals have faced sustained governmental pressure to reduce the number of managers and to limit their remuneration. In such settings, where management is perceived to be more an administrative than an entrepreneurial function, it is unclear how much management matters.

There is a large body of work exploring the contribution of management to the performance of private sector organisations (Bloom and Van Reenen, 2011) and (Bender et al., 2018). Private sector managers are employed by owners or shareholders primarily, if not exclusively, to improve profitability. Private sector organisations wishing to do better seek to attract better managers, offering them generous financial incentives. Senior managers are well-paid, and often there is a large pay gap with other staff in the organisation. Larger incentives are assumed to both attract better managers and, if appropriately structured, induce them to work harder (Murphy, 1999). Managers are granted authority over staff, allowed to make hiring and firing decisions, given considerable autonomy in their day-to-day decision making, and encouraged to take risks if these bring the promise of future financial rewards (Ichniowski et al., 1997).

The role of NHS hospital managers is quite different to those in the private sector. For one thing, there are not very many hospital managers. Approximately 3% of staff in NHS hospitals are employed in management roles as compared to 11% of staff employed in management roles in the economy overall in England (Office for National Statistics, 2018). Nor are NHS managers particularly well-paid compared to those working in the private sector or relative to the other staff in their organisation (Janke et al., 2018).

Moreover, while managing NHS hospitals is a complex undertaking, managers are more circumscribed in their role than private sector counterparts. NHS hospitals do not have owners or shareholders, but serve the interests of multiple stakeholders, and typically are not driven to maximise profits, but rather have to balance the pursuit of multiple, perhaps conflicting, objectives (Dixit, 2002) and (Burgess and Ratto, 2003). There is some international evidence that managers have limited authority within the hospital where clinicians make most of the key decisions about what the hospital does and how it goes about doing it, with managerial influence relying less on authority and more on persuasion (Harris, 1977). Risk-taking is discouraged, the managerial task being to ensure that things run smoothly. Health care is heavily regulated to maintain quality standards and financial fiduciary with hospitals carrying an associated bureaucratic burden in administering adherence to these regulations. These administrative tasks are separate from and, on a day-to-day basis subordinate to, clinical activities. Generally, the more critical are clinical activities, the more administrative management is confined, in terms of the bounds of its authoritative scope. It may be that NHS management is largely confined to these administrative roles, ensuring regulatory standards and requirements are met rather than inherently improving performance.

In this context, the relationship between the degree of managerial input and performance is likely to be tenuous at best, if evident at all. Previous studies in this area have modelled hospital output as a function of capital, physicians and management input. Management input has been conceived either as a distinct staff group in the hospital production function, as in Street et al. (1999) and Soderlund (1999), or as a way to influence total factor productivity, as in Bloom et al. (2016). Thus far these approaches have found very little association between the degree of management input and hospital output. This conclusion also seems to be borne out by the limited literature exploring the relationship between managerial pay and hospital performance. In a study of non-profit US hospitals Joynt et al. (2014) found no correlation between chief executive officer (CEO) compensation and patient outcomes. In their study of English NHS hospitals Janke et al. (2018) found that CEOs, regardless of their level of remuneration, appeared to have no discernible effect on any of a wide range of hospital performance measures.

Other studies have attempted to assess the quality of hospital management, and have found this to be related positively to hospital performance. As part of their work on global management structures Bloom et al. (2012) surveyed clinical practice leads in cardiology and orthopaedic departments in hospitals across several countries and found an association between higher quality management and lower risk-adjusted hospital mortality from acute myocardial infraction (AMI). A similar conclusion was reached by Bloom et al. (2015) where, based on interviews of managers in orthopaedic and cardiology departments within UK hospitals, higher management scores were associated with improved AMI survival rates, improved survival rates from general surgery, shorter waiting times and lower staff turnover.

In this paper we assess whether there is an association between managerial input and hospital performance in the English NHS. In order to do so we create measures of the amount and quality of managerial input as well as a collection of indicators against which to assess hospital performance. We advance the existing literature in four main ways. First, we exploit a relatively new workforce dataset that identifies the type and amount of staffing input used by all English NHS hospitals. Recent NHS studies drew upon the annual workforce census (WC), that took an annual snapshot of those working in the NHS. We utilise the monthly electronic staff record (ESR), an integrated human resources and payroll system that contains data for 1.4 million NHS employees. The ESR is comprehensive, capturing everybody on the payroll, and is highly granular, with non-medical staff categorised into 538 separate groups, each of which can be further split by pay-grade. We use the ESR to derive an accurate measure of the number of managers employed by NHS hospitals.

Second, we measure the quality of management by drawing upon information in the NHS staff survey. This is the largest workforce survey in the world, built on the responses from approximately 500,000 staff per year, in which staff answer a series of questions about the quality of management in their organisation.

Third, we do not confine our analysis to a single measure of hospital performance. Instead, recognising that NHS hospitals are tasked to pursue multiple objectives, we consider five indicators, capturing financial performance and measures of timely and high quality care.

Finally, where previous studies in this area have tended to analyse cross-sectional data, we construct and analyse a panel data set for all 129 English acute non-specialist NHS hospitals (termed “NHS trusts”) covering seven years from 2012/13 to 2018/19. The dataset contains a heterogeneous group of hospitals varying in a range of dimensions not directly captured in the data. We use our panel data together with hospital fixed effects to control for this unobserved heterogeneity.

In the remainder of this article we outline a suggested mechanism linking management input to outcomes, define our measures of management quantity and management quality and describe how we construct these measures, introduce our empirical specification, report our key results and conclude with a discussion and interpretation of our findings.

## 2 Suggested mechanism

The role of management in NHS hospitals can be broadly categorised into three related activities, management of staff, management of budgets and management of patients. As depicted in figure 1, the relationship between the quantity of managers and outcomes can be seen to be mediated by management quality. If a hospital can increase the quantity of management input, this would result in each manager managing fewer staff, a smaller budget and fewer beds. The closer management of these resources should, if more management is effective, result in these resources being better managed and thereby producing better outcomes for the hospital. This increase in management productivity may also be achieved in other ways such as by hiring better quality or more experienced managers rather than simply by hiring more managers.

**Figure 1:**
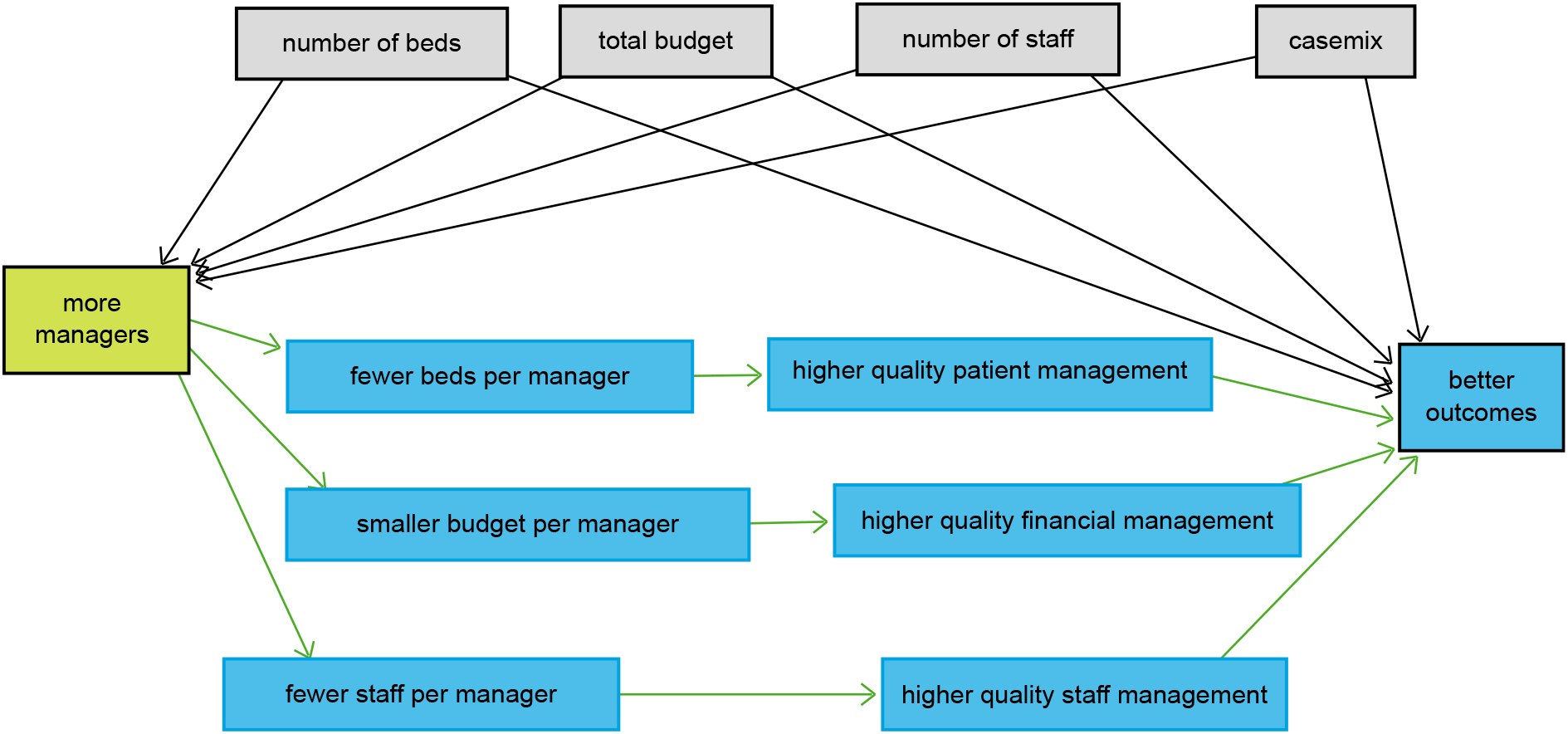
Suggested mechanism linking management input and hospital performance

The relationship between the quantity and quality of management might be stronger in private sector organisations than in public sector hospitals where the role of management is more focused on administration. We will, therefore, test part of the proposed mechanism linking increased management input to outcomes as mediated by higher quality staff management using data from the NHS staff survey. Whilst higher quality management of staff resulting in a more motivated and productive workforce will be crucial to improvement on all hospital outcomes, higher quality patient management might be particularly important in improving patient throughput and waiting times outcomes whilst higher quality financial management might be more important in managing the overall financial position of the hospital.

Managers in NHS hospitals, however, work under a number of constraints and have little influence over the total number of staff working at the hospital, the total budget envelope the hospital has to operate within, the total number of beds they can provide for patients, or the types of patients that receive treatment. These can be considered exogenous constraints that potentially confound the relationship between the number of managers and hospital outcomes. We will return to the relationships described in figure 1 to construct our empirical specification.

## 3 Data

We construct a panel data set consisting of all 129 non-specialist acute hospitals in the English NHS covering the seven financial years 2012/13 through to 2018/19. A full list of data sources used to construct our data set and a list of the hospitals included in the panel are detailed in tables 1, 2 and 3 of supplementary appendix.

**Table 1:**
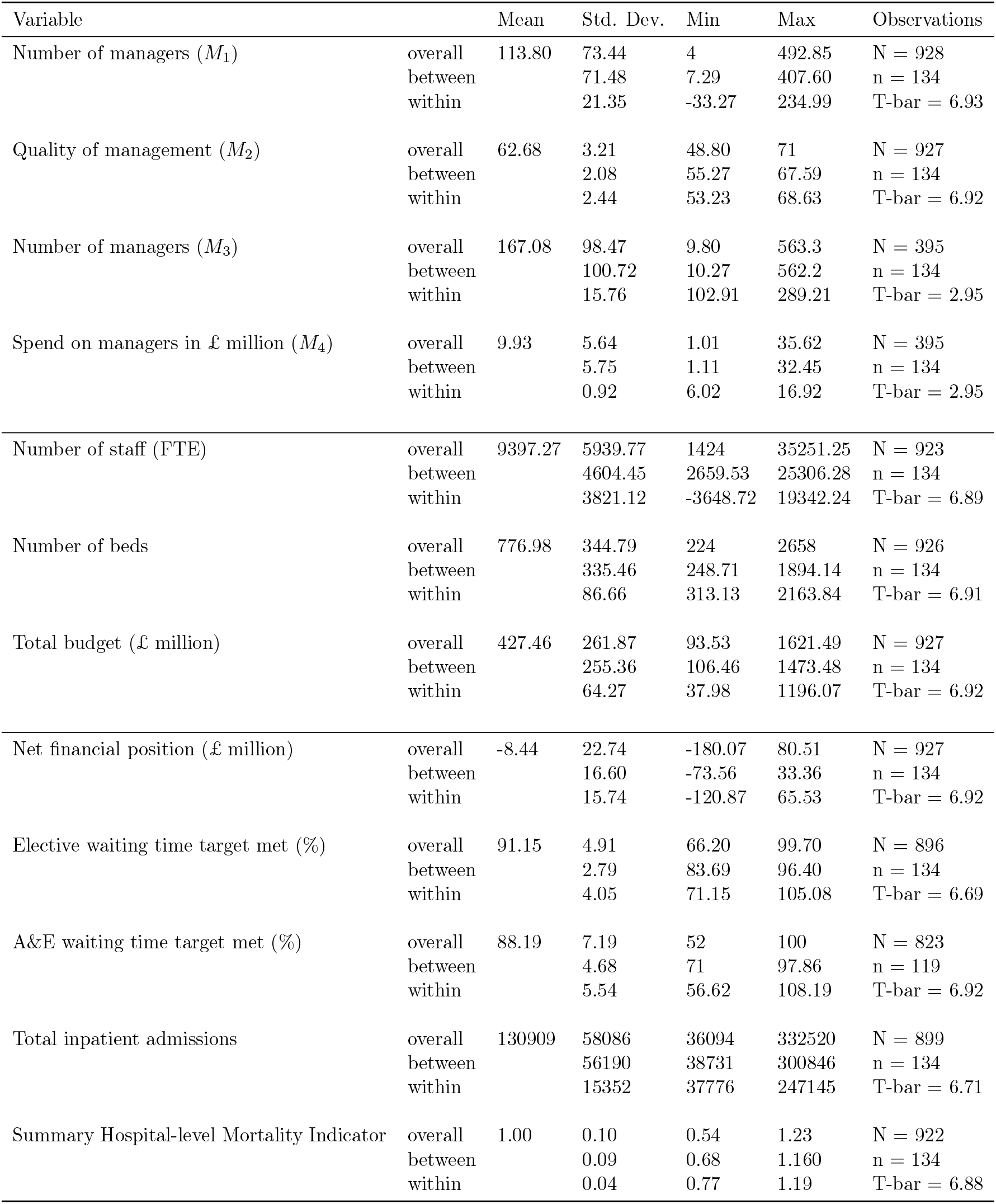
Overall, between and within variation in management, control and outcome variables 2012/13 – 2018/19

**Table 2:**
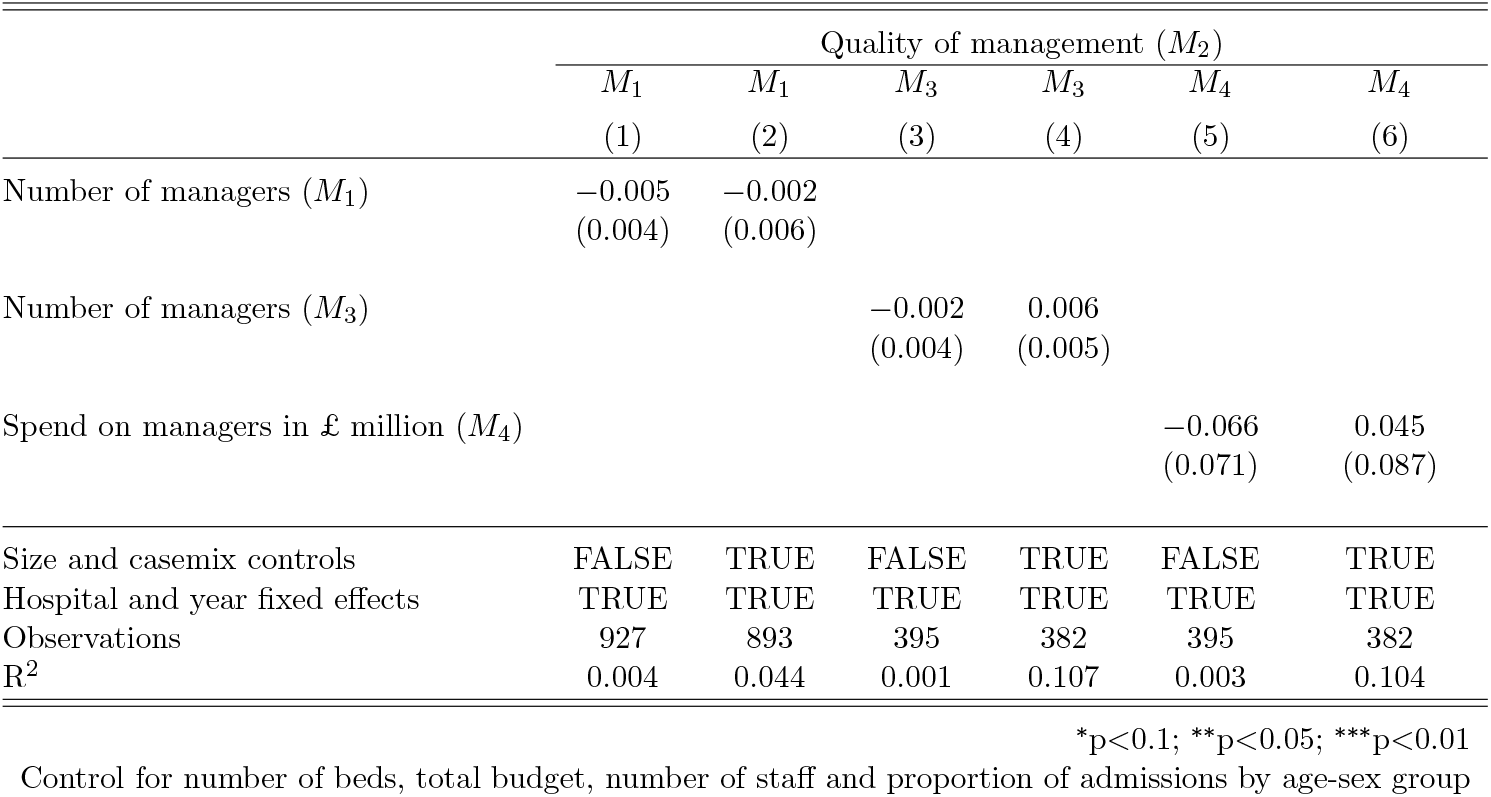
Relationship between quantity of managerial input and managerial quality 2012/13 – 2018/19

| | Quality of management ($M_2$) | | | | | |
| --- | --- | --- | --- | --- | --- | --- |
| | $M_1$ | $M_1$ | $M_3$ | $M_3$ | $M_4$ | $M_4$ |
|  | (1) | (2) | (3) | (4) | (5) | (6) |
| Number of managers ($M_1$) | -0.005<br>(0.004) | -0.002<br>(0.006) | | | | |
| Number of managers ($M_3$) | | | -0.002<br>(0.004) | 0.006<br>(0.005) | | |
| Spend on managers in £ million ($M_4$) | | | | | -0.066<br>(0.071) | 0.045<br>(0.087) |
| Size and casemix controls | FALSE | TRUE | FALSE | TRUE | FALSE | TRUE |
| Hospital and year fixed effects | TRUE | TRUE | TRUE | TRUE | TRUE | TRUE |
| Observations | 927 | 893 | 395 | 382 | 395 | 382 |
| R<sup>2</sup> | 0.004 | 0.044 | 0.001 | 0.107 | 0.003 | 0.104 |

\*p<0.1; \*\*p<0.05; \*\*\*p<0.01
Control for number of beds, total budget, number of staff and proportion of admissions by age-sex group

**Table 3:** Amount of managerial input and Net financial position (£ million) 2012/13 – 2018/19

|  | Net financial position (£ million) |  |  |  |  |  |  |  |
| --- | --- | --- | --- | --- | --- | --- | --- | --- |
| | $M_1$ | $M_1$ | $M_2$ | $M_2$ | $M_3$ | $M_3$ | $M_4$ | $M_4$ |
|  | (1) | (2) | (3) | (4) | (5) | (6) | (7) | (8) |
| Number of managers ($M_1$) | -0.007<br>(0.057) | -0.011<br>(0.047) | | | | | | |
| Quality of management ($M_2$) | | | 0.835<br>(0.561) | 0.433<br>(0.441) | | | | |
| Number of managers ($M_3$) | | | | | -0.054<br>(0.088) | 0.124<br>(0.120) | | |
| Spend on managers in £ million ($M_4$) | | | | | | | -0.916<br>(1.564) | 2.022<br>(2.345) |
| Size and casemix controls | FALSE | TRUE | FALSE | TRUE | FALSE | TRUE | FALSE | TRUE |
| Hospital and year fixed effects | TRUE | TRUE | TRUE | TRUE | TRUE | TRUE | TRUE | TRUE |
| Observations | 927 | 893 | 927 | 893 | 395 | 382 | 395 | 382 |
| R<sup>2</sup> | 0.0001 | 0.032 | 0.008 | 0.034 | 0.004 | 0.062 | 0.004 | 0.062 |

\*p<0.1; \*\*p<0.05; \*\*\*p<0.01
Control for number of beds, total budget, number of staff and proportion of admissions by age-sex group

### 3.1 Quantifying managerial input

There is no agreed measure of managerial input and it is a challenge to establish how best to assess the form, quantity and quality of hospital management and to differentiate general administrative functions from clinical management. To meet this challenge, we define four different measures of managerial input, the first two of which can be constructed for the full period, the other two just for the most recent three years.

The first measure, referred to as *M*_1_, can be constructed for all years in the series. This is a simple count of the number of full-time equivalent (FTE) managers in each hospital, using data from the NHS electronic staff records (ESR) which is part of the information system used to manage human resources in the NHS. The monthly data published from the ESR captures staff role and FTEs of all permanent NHS staff, including managers and senior managers. We identify managers as those staff in post in September, midway through the financial year, having the word “manager” or “senior manager” as their primary job title. Hospitals with more managers might be better managed with correspondingly higher performance.

The second measure, *M*_2_, captures managerial quality, using data from the NHS staff survey. This is the largest workforce survey in the world and has been conducted every year since 2003. In the 2018/19 wave of the staff survey around 1.1 million NHS employees were invited to complete the survey of which some 500,000 responded (a response rate of 46%) (Staff Survey Coordination Centre, 2018). Responses were scored on a scale from 0 to 4 corresponding to answers: Strongly disagree (0), Disagree (1), Neither agree nor disagree (2), Agree (3), Strongly agree (4). There were 11 questions on the staff survey that focused specifically on management – 7 on immediate managers and 4 on senior managers (see supplementary appendix table 2 for the full list of these questions). We normalised the score for each of the 11 questions on a percentage scale and then averaged across all the questions to produce the *M*_2_ measure of managerial quality for each NHS hospital. As robustness checks, we also ran analyses using responses for each of the 11 questions separately. Hospitals with more highly regarded managers might have higher levels of performance.

From 2016/17 the ESR data were enriched by including more detailed job descriptions and pay grade information. Previously the ESR categorised non-medical staff into just 19 categories; from 2016/17 onward non-medical staff were categorised into 538 distinct roles. The use of the pay grade data made it possible to limit identification of non-medical staff with managerial roles to those on NHS pay scales of grade 7 or above (£31,383 in 2016/17), as those below this grade are likely to have largely administrative responsibilities (NHS Digital, 2018). This more precise definition thus includes nurse managers but excludes junior staff who are likely to be in administrative positions. Using this greater level of detail we were able to adopt a more precise definition of the number of managers referred to as *M*_3_. This definition also includes those staff likely to have senior managerial responsibilities but without having the word “manager” in their job title.

The enriched ESR also allowed us to construct our fourth measure of managerial input, referred to as *M*_4_, by weighting the number of FTEs at each managerial grade by the associated pay rate. Theoretically, pay should reflect the marginal product of labour, with higher paid managers receiving rewards commensurate with their contribution Murphy (1999). Hospitals that offer more generous pay packages would expect a greater performance return from their better paid managers.

In 2018/19 the ESR recorded a total of 732,038 FTE staff in the 129 acute NHS hospitals. Of these, 16,978 (2.1%) had manager or senior manager in their primary job title (*M*_1_) and 22,751 (3.1%) are identified as managers if we adopt our more precise definition (*M*_3_) to identify managers. An important caveat is that medical staff who may have management responsibility, such as senior doctors (known as “consultants” in the NHS) who constitute 5.3% of the workforce, are not included in these manager numbers. Both total numbers of staff and numbers of managers have steadily increased in the NHS hospitals we study over the seven years we analyse by 13% and 23% respectively (see supplementary appendix figure 1 for a description of trends over time).

The distributions of the four measures of managerial input across our sample of 129 non-specialist NHS acute hospitals in 2018/19 are shown in figure 2. Descriptive statistics and analytical results for the other years of data are given in section 2 of the supplementary appendix. There is variation between hospitals in both quantity and quality of managerial input. In 2018/19 managers made up between 0.26% and 7.78% of the hospital workforce and accounted for between 0.31% and 4.28% of total operating costs. The consolidated management score derived from the NHS staff survey ranges from 56.5% for the lowest scoring hospitals to above 70.8% for the highest scoring hospitals in our sample.

**Figure 2:**
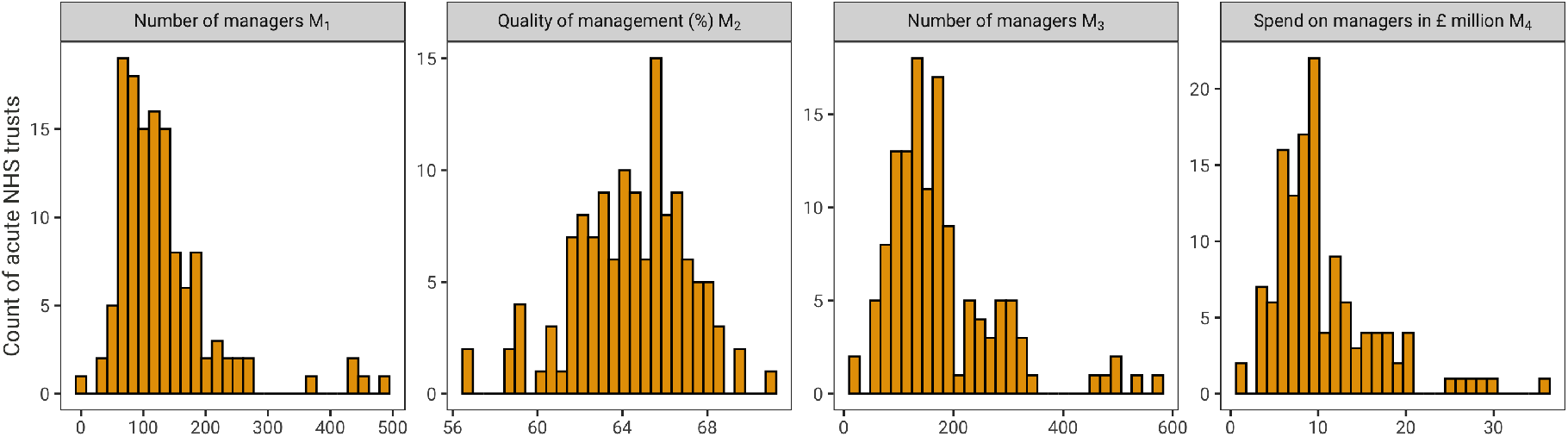
Distribution of management input variables across non-specialist acute NHS hospitals in 2018/19

### 3.2 Outcome measures

While it is generally accepted that private sector organisations are interested primarily in maximising profit, there is no single objective that not-for-profit or public sector organisations aim to achieve. Hospitals pursue multiple objectives, but there is no consensus on what these are. In our analysis, we measure hospital performance using five indicators. The first indicator is a measure of financial performance while the remainder capture the hospital’s mission to deliver timely and high quality care to those in need of treatment.

Financial performance is measured by *net financial position*, this indicator being derived from the hospital’s annual accounts. A positive value indicates that total hospital revenues exceed total operating costs. Timely care is captured by two indicators, both used by the English government to monitor hospital performance. The *elective waiting time* measures the proportion of patients who are treated within 18 weeks of being referred by their general practitioner for a planned hospital admission. The *A&E waiting time* measures the proportion of patients in accident and emergency (A&E) who are seen within 4 hours.

The last two indicators capture the quantity and quality of care that the hospital provides. The number of *inpatient admissions* measures the total number of patients (measured as “finished consultant episodes”) at each hospital. The *Summary Hospital-level Mortality Indicator* (SHMI) reports the ratio between the actual number of patients who die following hospitalisation and the number that would be expected to die on the basis of case mix adjusted average England figures. The SHMI is the official mortality indicator for acute hospitals in England, with values greater than one indicating worse than average performance.

The distribution of these five performance indicators across the 129 acute NHS hospitals in our sample for financial year 2018/19 are plotted in figure 3. There is variation between hospitals in each of the indicators, with evidence of poor performance: the majority of hospitals (67%) are in deficit; all hospitals fail to ensure that all of their patients are treated within 18 weeks of referral; most hospitals fall well below the target that 95% of A&E patients are seen within fours hours; and the SHMI is worse than expected for 52% of hospitals. Similar descriptions for the other years of data are given in section 2 of the supplementary appendix (figures 2-4).

**Figure 3:**
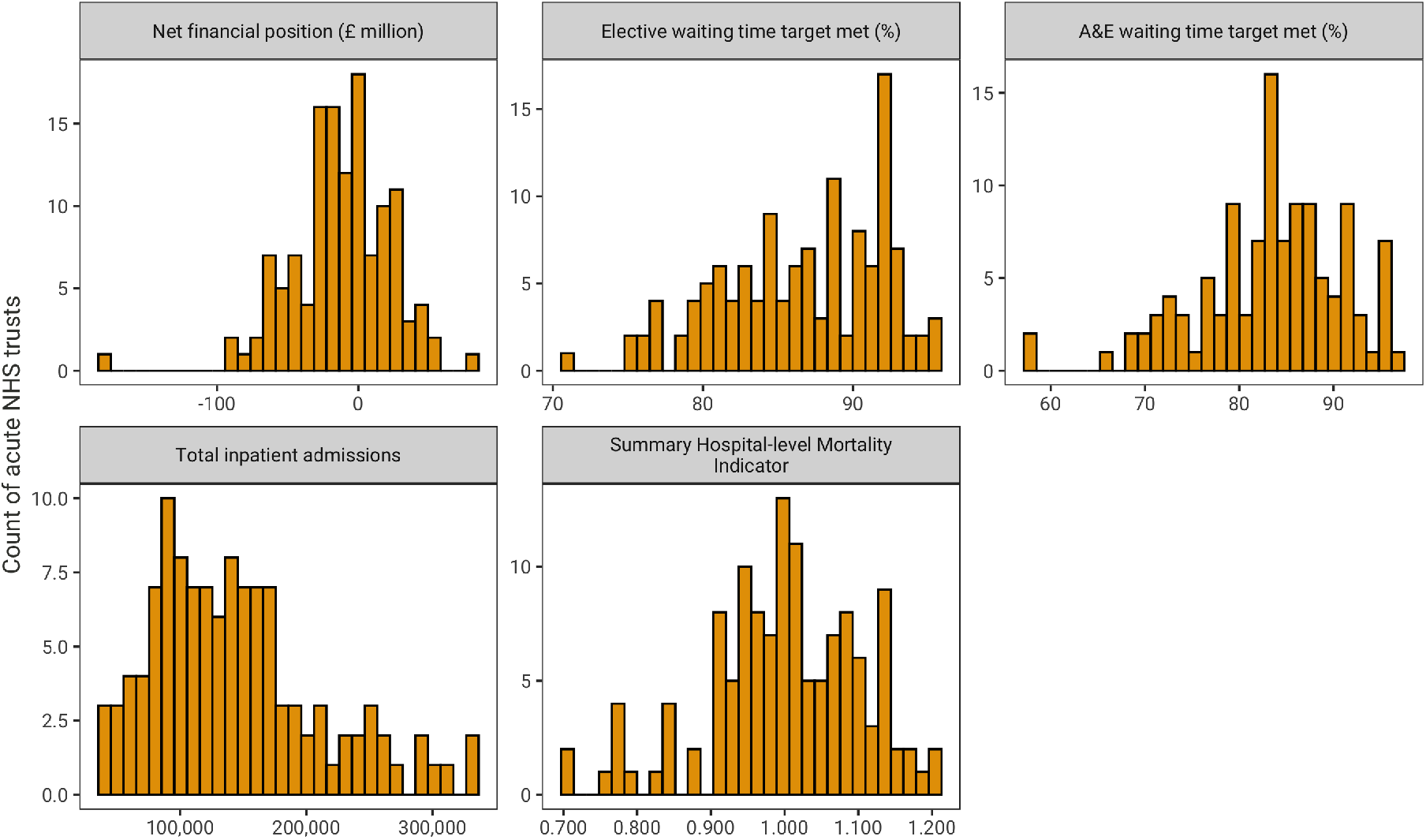
Distribution of hospital outcome variables across non-specialist acute NHS hospitals in 2018/19

### 3.3 Controls

The ability of management to influence performance may depend on the size of the hospital, if there are scale effects, and the diversity of activity, if there are scope effects. These are factors that managers in NHS hospitals have little power to influence so can be seen as constraints under which they operate. We capture differences in hospital size using three measures: the total number of beds in the hospital; the total budget (as measured by the hospital’s total operating cost); and the number of staff.

The distribution of these size variables are depicted in figure 4. The diversity of activity is captured by proportions of inpatient hospital admissions by age-group and sex, reflecting the hospital’s casemix.

**Figure 4:**
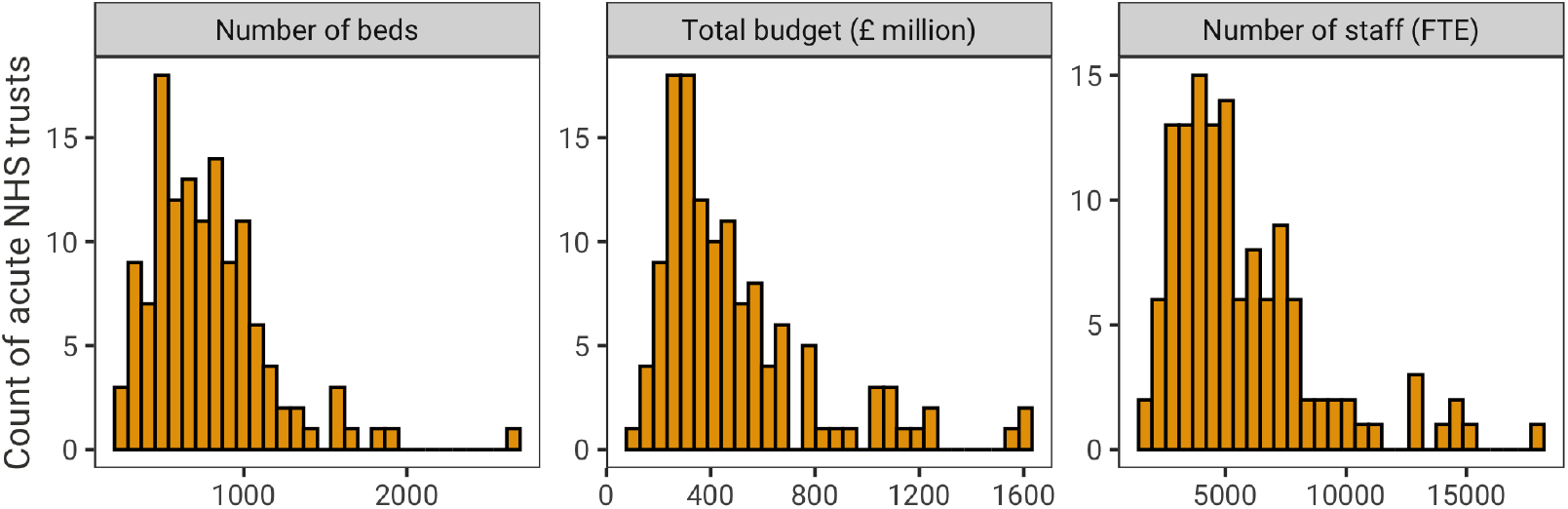
Distribution of control variables across non-specialist acute NHS hospitals in 2018/19

In the analysis we divide each of the size measures by the number of managers employed by the hospital to get an impression of how intensely these are managed. We plot these against the proportion of staff working in managerial positions at each hospital in Figure 5. The negative slopes in this figure indicate that as the proportion of staff working in managerial positions increases then, on average, each manager is responsible for fewer beds, fewer staff, and a decreasing amount of the hospital’s budget. This suggests that those hospitals employing more managers are managing these resources more closely than those with fewer managers. This ties back to our proposed mechanism depicted in figure 1 where more closely managed resources mediate the relationship between increased management input, better resource management, and better outcomes.

**Figure 5:**
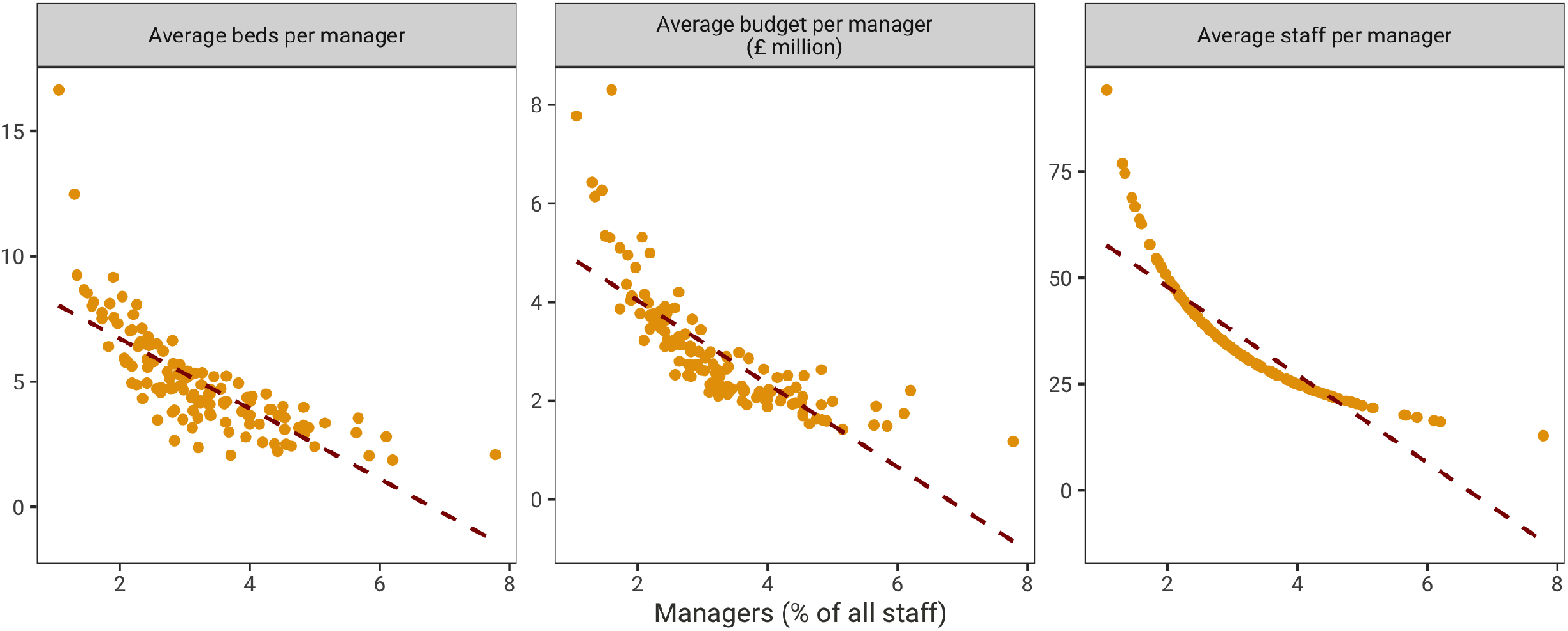
Relationship between management input and resources to be managed across non-specialist acute NHS hospitals in 2018/19

Similar figures for the other years in our data set are given in section 2 of the supplementary appendix.

### 3.4 Comparing the variation in the data set between hospitals and within hospitals over time

Descriptive statistics for the management input, hospital controls and hospital outcomes variables for the full seven years of the panel are given in table 1. With the exception of quality of management, *M*_2_, there is substantial variation in all variables and, in general, variation between hospitals is larger than variation within hospitals over time. This variation is confirmed in the supplementary appendix figures 12 and 13 which plot the relationship between management input and each of the hospital outcome variables between hospitals (figure 12) and within hospitals over time (figure 13).

## 4 Empirical specification

### 4.1 Does the quantity of managerial input impact management quality?

We first assess whether the quantity of managerial input, measured as either *M*_1_, *M*_3_ or *M*_4_, is associated with staff perceptions of managerial quality, *M*_2_, as measured by the combined responses to the 11 management related questions on the NHS staff survey. The hypothesis is that having more managers might impact outcomes, at least in part, by improving the overall quality of staff management (see figure 1). If so, we would go on to estimate the degree to which the relationship between quantity of management and hospital outcomes is mediated by the impact of quantity of management on perceived management quality. If not, *M*_2_ can be considered an alternative measure of managerial input in its own right.

We assess this association by estimating the following equation in which the set of three measures of managerial quantity *M*_*Q*_(*M*_1_, *M*_3_, *M*_4_) are entered alternatively:

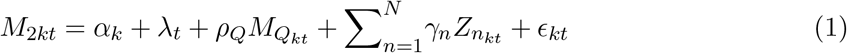

where *k* indexes the hospitals, *ρ*_*Q*_ are the parameters on managerial quantity to be estimated, *α* are the hospital fixed effects, *λ* are the year fixed effects, *γ* are the parameters relating to the time varying size and casemix control variables, and *E* is a standard fixed effects model error term. This equation is estimated for the full data series, 2012/13 to 2018/19, when using the *M*_1_ measures, and for years 2016/17 through 2018/19, when using the *M*_3_ and *M*_4_ measures. All equations are estimated with and without controls.

Hospitals in the NHS differ in a host of other dimensions including the number and range of clinical services that they provide and how they divide their physical infrastructure amongst these services. We are unable to capture detailed information about such dimensions from the publicly available data and so include hospital fixed effects *α*_*k*_ in our empirical specification to account for this time invariant unobserved heterogeneity between the hospitals. Hausman tests indicate that fixed effects are preferred to random effects in 2 out of 3 of these equations. In section 4 of the supplementary index, we report pooled, between, random effects and lagged dependent variables specifications as robustness checks.

We also include year fixed effects *λ*_*t*_ to account for period specific effects and general time trends that would impact all hospitals in our sample. We adjust the standard errors of our estimates to account for the hierarchical clustering of our data.

We additionally run our model separately on responses to each of the 11 individual questions regarding management in the NHS staff survey in place of the overall management quality measure *M*_2_ constructed from combining the responses.

### 4.2 Does managerial input impact hospital outcomes?

We then analyse the relationship between managerial input and each of our outcomes *y* again using a fixed effects ordinary least squares regression. We consider in turn our four measures of managerial input *M*_*_(*M*_1_ … *M*_4_), each entered as a linear term, with the equation specified as:

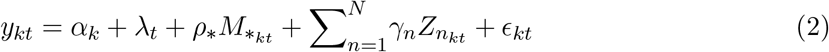

where all terms are specified as for equation 1, but with *ρ*_*_ being the parameters on managerial input to be estimated. As previously, we use Hausman tests to validate our choice of fixed effects, this being preferred to random effects in 18 of the 20 equations. For completeness we report pooled, between, random effects and lagged dependent variables specifications as robustness checks in section 6 of supplementary index. We also re-run our analysis separately substituting *M*_2_ for each of the 11 staff survey questions.

We conducted the analysis in the R statistical software (version 4.0.3) using the plm package (version 2.4-1). All data used in the analysis along with the analysis code is available at the following url: https://github.com/miqdadasaria/hospital_management

## 5 Results

### 5.1 Does the quantity of managerial input impact management quality?

The results of our fixed effects regressions of equation 1 examining the relationship between the quantity and quality of managerial input with and without controls are given in table 2. We find no evidence of a significant relationship between the quantity of management, whether measured as *M*_1_, *M*_3_ or *M*_4_, on management quality.

Looking at alternative specification of these relationships in tables 5-7 of the supplementary appendix we find a significant relationship only in the pooled models where there is evidence of a positive relationship between each of the three quantity measures and quality. However, the pooled model is ruled out using a Breusch-Pagan test.

The results where *M*_2_ is replaced with each of the 11 questions in the staff survey are given in tables 8-10 of the supplementary appendix. Consistent with the findings for their impact on *M*_2_ none of the three measures of quantity of management are found to significantly impact any of the responses to these individual questions in the fixed effects model specification.

### 5.2 Does managerial input impact hospital outcomes?

Results from estimating equation 2 are reported for each of the five outcomes in tables 3 to 7. Each table reports the estimates for each measure of managerial input, for equations with and without controls. There are no significant associations between any of the measures of managerial input and *net financial position* (Table 3), the *proportion of elective patients who are treated within 18 weeks of being referred* (Table 4), the *proportion of A&E patients seen within 4 hours* (Table 5) and the *SHMI* (Table 7). There is a positive association between the *number of admissions* and the number of managers and a negative association with managerial quality (Table 6), but these associations are significant only in the absence of the set of size and casemix control variables.

**Table 4:** Amount of managerial input and Elective waiting time target met (%) 2012/13 – 2018/19

|  | Elective waiting time target met (%) |  |  |  |  |  |  |  |
| --- | --- | --- | --- | --- | --- | --- | --- | --- |
| | $M_1$ | $M_1$ | $M_2$ | $M_2$ | $M_3$ | $M_3$ | $M_4$ | $M_4$ |
|  | (1) | (2) | (3) | (4) | (5) | (6) | (7) | (8) |
| Number of managers ($M_1$) | 0.006<br>(0.006) | 0.011<br>(0.007) | | | | | | |
| Quality of management ($M_2$) | | | 0.024<br>(0.097) | −0.020<br>(0.093) | | | | |
| Number of managers ($M_3$) | | | | | 0.003<br>(0.011) | 0.013<br>(0.017) | | |
| Spend on managers in £ million ($M_4$) | | | | | | | 0.098<br>(0.188) | 0.317<br>(0.275) |
| Size and casemix controls | FALSE | TRUE | FALSE | TRUE | FALSE | TRUE | FALSE | TRUE |
| Hospital and year fixed effects | TRUE | TRUE | TRUE | TRUE | TRUE | TRUE | TRUE | TRUE |
| Observations | 896 | 865 | 896 | 865 | 384 | 371 | 384 | 371 |
| R<sup>2</sup> | 0.002 | 0.021 | 0.0002 | 0.018 | 0.0004 | 0.067 | 0.001 | 0.069 |

\*p<0.1; \*\*p<0.05; \*\*\*p<0.01
Control for number of beds, total budget, number of staff and proportion of admissions by age-sex group

**Table 5:** Amount of managerial input and A&E waiting time target met (%) 2012/13 – 2018/19

|  | A&E waiting time target met (%) |  |  |  |  |  |  |  |
| --- | --- | --- | --- | --- | --- | --- | --- | --- |
| | $M_1$ | $M_1$ | $M_2$ | $M_2$ | $M_3$ | $M_3$ | $M_4$ | $M_4$ |
|  | (1) | (2) | (3) | (4) | (5) | (6) | (7) | (8) |
| Number of managers ($M_1$) | −0.013<br>(0.010) | −0.016<br>(0.010) | | | | | | |
| Quality of management ($M_2$) | | | 0.110<br>(0.102) | 0.122<br>(0.107) | | | | |
| Number of managers ($M_3$) | | | | | 0.004<br>(0.018) | −0.023<br>(0.024) | | |
| Spend on managers in £ million ($M_4$) | | | | | | | −0.042<br>(0.322) | −0.604<br>(0.417) |
| Size and casemix controls | FALSE | TRUE | FALSE | TRUE | FALSE | TRUE | FALSE | TRUE |
| Hospital and year fixed effects | TRUE | TRUE | TRUE | TRUE | TRUE | TRUE | TRUE | TRUE |
| Observations | 823 | 817 | 823 | 817 | 353 | 353 | 353 | 353 |
| R<sup>2</sup> | 0.004 | 0.034 | 0.002 | 0.033 | 0.0003 | 0.054 | 0.0001 | 0.062 |

\*p<0.1; \*\*p<0.05; \*\*\*p<0.01
Control for number of beds, total budget, number of staff and proportion of admissions by age-sex group

**Table 6:**
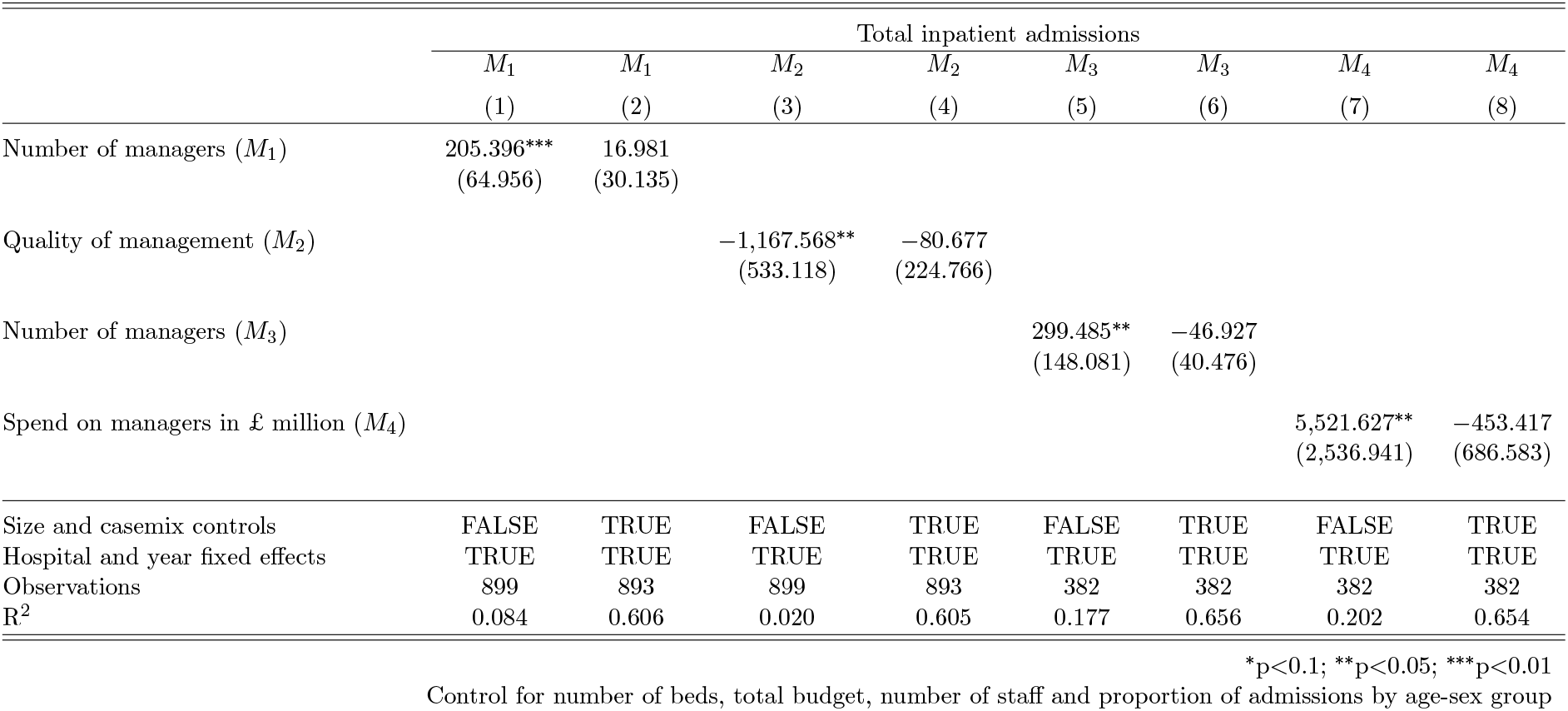
Amount of managerial input and Total inpatient admissions 2012/13 – 2018/19

|  | Total inpatient admissions |  |  |  |  |  |  |  |
| --- | --- | --- | --- | --- | --- | --- | --- | --- |
| | $M_1$ | $M_1$ | $M_2$ | $M_2$ | $M_3$ | $M_3$ | $M_4$ | $M_4$ |
|  | (1) | (2) | (3) | (4) | (5) | (6) | (7) | (8) |
| Number of managers ($M_1$) | 205.396***<br>(64.956) | 16.981<br>(30.135) | | | | | | |
| Quality of management ($M_2$) | | | -1,167.568**<br>(533.118) | -80.677<br>(224.766) | | | | |
| Number of managers ($M_3$) | | | | | 299.485**<br>(148.081) | -46.927<br>(40.476) | | |
| Spend on managers in £ million ($M_4$) | | | | | | | 5,521.627**<br>(2,536.941) | -453.417<br>(686.583) |
| Size and casemix controls | FALSE | TRUE | FALSE | TRUE | FALSE | TRUE | FALSE | TRUE |
| Hospital and year fixed effects | TRUE | TRUE | TRUE | TRUE | TRUE | TRUE | TRUE | TRUE |
| Observations | 899 | 893 | 899 | 893 | 382 | 382 | 382 | 382 |
| R<sup>2</sup> | 0.084 | 0.606 | 0.020 | 0.605 | 0.177 | 0.656 | 0.202 | 0.654 |

\*p<0.1; \*\*p<0.05; \*\*\*p<0.01
Control for number of beds, total budget, number of staff and proportion of admissions by age-sex group

**Table 7:** Amount of managerial input and Summary Hospital-level Mortality Indicator 2012/13 – 2018/19

|  | Summary Hospital-level Mortality Indicator |  |  |  |  |  |  |  |
| --- | --- | --- | --- | --- | --- | --- | --- | --- |
| | $M_1$ | $M_1$ | $M_2$ | $M_2$ | $M_3$ | $M_3$ | $M_4$ | $M_4$ |
|  | (1) | (2) | (3) | (4) | (5) | (6) | (7) | (8) |
| Number of managers ($M_1$) | -0.00002<br>(0.0001) | 0.00000<br>(0.0001) | | | | | | |
| Quality of management ($M_2$) | | | -0.0001<br>(0.001) | 0.0001<br>(0.001) | | | | |
| Number of managers ($M_3$) | | | | | -0.0001<br>(0.0001) | -0.0003<br>(0.0002) | | |
| Spend on managers in £ million ($M_4$) | | | | | | | -0.002<br>(0.002) | -0.005<br>(0.004) |
| Size and casemix controls | FALSE | TRUE | FALSE | TRUE | FALSE | TRUE | FALSE | TRUE |
| Hospital and year fixed effects | TRUE | TRUE | TRUE | TRUE | TRUE | TRUE | TRUE | TRUE |
| Observations | 922 | 888 | 922 | 888 | 390 | 377 | 390 | 377 |
| R<sup>2</sup> | 0.0001 | 0.034 | 0.00002 | 0.034 | 0.002 | 0.040 | 0.002 | 0.040 |

\*p<0.1; \*\*p<0.05; \*\*\*p<0.01
Control for number of beds, total budget, number of staff and proportion of admissions by age-sex group

The results of the various ways of accounting for between and within hospital variation in the data are presented in tables 11-30 of the supplementary appendix. These are ordered to show, for each outcome in turn, four tables corresponding to each measure of managerial input. When the control variables are included the association between the outcomes and the three measures of the quantity of managerial input, *M*_1_, *M*_3_ and *M*_4_, remains insignificant across these alternative specifications.

However, in contrast to the fixed effects model, under these alternative models the management quality variable *M*_2_ has a significantly positive association with better performance on *net financial position* (supplementary appendix table 12), the *proportion of elective patients who are treated within 18 weeks of being referred* (supplementary appendix table 16), and the *proportion of A&E patients seen within 4 hours* (supplementary appendix table 20). Whilst these alternative specifications are ruled out by the Breusch-Pagan and Hausman tests, it is worth noting that variation in the *M*_2_ measure within hospitals over time was modest (see table 1) making it difficult for the fixed effects model to identify any relationship. In light of the results of the robustness tests and lack of variation in management quality over time, we conclude that there is likely a positive relationship between management quality and outcomes that we are unable to pick up in our fixed effects model.

Finally, the results for the fixed effects model where *M*_2_ is replaced with each of the questions in the staff survey are given in tables 31-35 of the supplementary appendix. Consistent with the findings for *M*_2_ none of the responses to these individual questions are significantly associated with changes in hospital outcomes under the fixed effects model specification.

## 6 Discussion

Our suggested mechanism linking the quantity of management input to hospital performance set out in figure 1 posited that, if a hospital can increase the quantity of management input, it should be in a better position to provide higher quality management of staff, finances, and patients and, consequently, produce better outcomes for the hospital.

We find no evidence of an association between our measures of quantity of managerial input and quality of management as measured by the responses to the 11 management related questions on the NHS staff survey (*M*_2_). Furthermore, we find no associations between our measures of quantity of management input and five measures of hospital performance. This holds irrespective of how we define managerial input, whether by number of managers or expenditure on management. These results are generally robust to how we account for variation between hospitals and within hospitals over time. We conclude that hospitals hiring more managers neither see an improvement in the quality of management leading to better performance, nor does increasing the numbers of managers appear to improve hospital performance through any other direct or indirect mechanism.

Our data set does suggest a positive association between better management quality and hospital performance particularly on measures of financial performance, and elective and A&E waiting times. However, we have very little variation in our management quality measure within hospitals over time and these associations are not found to be significant under our default fixed effects model specification.

One plausible reason for the lack of an association between quantity of management input and hospital performance is that, compared to their private sector counterparts, many NHS managers have limited discretion in how they perform their roles. They are required to implement official instructions and to meet externally imposed targets. In this sense, NHS hospital management is largely about the administration of compliance to regulations and externally imposed national policies. Management reduces to administrative tasks to ensure that minimum standards of clinical and financial care, however defined, are not disregarded or breached. This is very different to the role of management in the private sector where managers typically have more autonomy, and enjoy considerable authority in deciding key objectives and in optimising their firm’s production processes. So, where the quantity of managerial input does vary in NHS hospitals over time it is for reasons unrelated to performance. These reasons include changes in the number of sites across which the hospital is spread and changes in the range and types of services offered to patients. The number of managers is largely determined by the administrative tasks that need to be fulfilled, with the scope of management circumscribed to these well defined tasks. Similarly, there is limited variation in the remuneration of NHS managers, hospitals being constrained in what pay packages they can offer. These factors leave little room for exceptionally good managers to shine or exceptionally bad managers to do much damage to overall hospital performance. Given the above considerations and after controlling for both observed and unobserved heterogeneity across hospitals, the lack of a systematic association between levels of management input and hospital performance is not surprising.

Our study has a number of strengths compared to previous studies on the subject. Firstly, we make use of the ESR, which captures payroll data on everybody employed by the NHS, and from which we are able to gain an accurate assessment of how many people were employed as managers and what pay scale they were on. Accuracy improved in the later years, when job roles in the ESR were reported in much greater detail.

Secondly, we capture the quality of management practice based on the NHS staff survey which includes responses of approximately 500,000 staff per year - the largest workforce survey in the world. Previous work in this area has measured the quality of management practices in a single year using telephone interviews with small numbers of staff in management positions within hospitals (Bloom et al., 2015, 2012; Tsai et al., 2015). Our use of this large survey allows us to build a panel of data on management practices as interpreted by a much wider range of staff and to capture changes over time within hospitals.

Thirdly, we analyse seven years of data for a sample of 129 non-specialist acute NHS hospitals. Hospitals in the NHS are hugely heterogeneous, differing in a host of dimensions including the number and range of clinical services that they provide and how they divide their physical infrastructure amongst these services. Given that we are unable to capture detailed information about such dimensions from the publicly available data, and that most of these hospital characteristics remain fairly constant over time, we include hospital fixed effects in our empirical models to control for the unobserved heterogeneity between hospitals. In doing so we necessarily limit our focus to examining how changes in numbers of managers within hospitals over time relate to changes in outcomes for these hospitals. Whilst this choice is supported by various statistical tests, we also explore the robustness of our conclusions to a range of alternative specifications including pooled, between effects, random effects and models including lagged dependent variables.

Finally, we use SHMI as our main clinical outcome variable - covering mortality across all specialities in the hospital. Previous studies have focused solely on acute myocardial infarction deaths, giving a partial view of this aspect of clinical performance see Bloom et al. (2015).

Of course our study has limitations. Firstly, the data do not detail the extent to which senior doctors are involved in management, so these doctors have been omitted from the management measures we construct. Secondly, whilst our empirical approach controls for unobserved time invariant heterogeneity between hospitals it is not able to capture the effect of any unobserved time varying heterogeneity between hospitals. Such effects may arise if unobserved hospital characteristics influence changes in the number and type of managers that are employed. For example additional managers may be appointed to help deliver service expansions.

Previous studies of the impact of quality of management on hospital performance find a positive relationship between better management practices and better hospital performance (Tsai et al., 2015; Bloom et al., 2015, 2012). Some of our analyses also suggest a positive relationship between quality of management and hospital performance, but in the fixed effects analyses these effects disappear. This suggests that either these management practices are largely time invariant and so swept out by the hospital fixed effects or that they are confounded by other time invariant unobserved factors in the cross-sectional analyses that the hospital fixed effects account for. Either way, there is no evidence of an impact of quantity of management input on management quality.

Our findings are consistent with previous studies that find little evidence of a clear positive relationship between the quantity of managerial input and hospital performance in the NHS (Street et al., 1999; Soderlund, 1999). Similarly, to these previous studies our findings provides no grounds either for reducing or increasing managerial input in NHS hospitals but raise the question of whether these managers are granted sufficient autonomy to make a difference.

## Data Availability

All data used in the manuscript are publicly available administrative datasets.

## Notes

### Competing Interest Statement

The authors have declared no competing interest.

### Clinical Trial

The study is based on administrative data

### Funding Statement

This work was funded by a grant from The Health Foundation

### Author Declarations

Ethics approval was not required for this study

