## Supplementary appendix for "The Impact of Management on Hospital Performance"

### 1 Data

#### 1.1 Data sources

Table 1: Data Sources

| Data | Source | URL |
|---|---|---|
| Inpatient admissions | NHS Digital | https://digital.nhs.uk/data-and-information/publications/statistical/hospital-admitted-patient-care-activity/2018-19 |
| Accident and emergency target | NHS Digital | https://digital.nhs.uk/data-and-information/publications/statistical/hospital-accident–emergency-activity/2018-19 |
| Inpatient admissions referral to treatment target | NHS England | https://www.england.nhs.uk/statistics/statistical-work-areas/rtt-waiting-times/rtt-data-2018-19/ |
| NHS staff survey management scores | NHS survey coordination centre | https://www.nhsstaffsurveys.com/Page/1064/Latest-Results/2018-Results/ |
| NHS workforce statistics (ESR) | NHS Digital | https://digital.nhs.uk/data-and-information/publications/statistical/nhs-workforce-statistics/september-2018 |
| Summary Hospital-level Mortality Indicator (SHMI) | NHS Digital | https://digital.nhs.uk/data-and-information/publications/clinical-indicators/shmi/ |
| Care quality commission rating | Care quality commission | https://www.cqc.org.uk/about-us/transparency/using-cqc-data |
| Financial performance of NHS trusts | NHS Improvement | https://improvement.nhs.uk/resources/nhs-providers-trust-accounts-consolidation-tac-data-201718/ |
| Bed Availability and Occupancy Data Ð Overnight | NHS England | https://www.england.nhs.uk/statistics/statistical-work-areas/bed-availability-and-occupancy/bed-data-overnight/ |

#### 1.2 Staff survey questions

Table 2: Staff Survey Management Questions

| **7) My immediate manager:** | **8) Senior managers:** |
|---|---|
| a. encourages those who work for her/him to work as a team | a. I know who the senior managers are here |
| b. can be counted on to help me with a difficult task at work | b. Communication between senior management and staff is effective |
| c. gives me clear feedback on my work | c. Senior managers here try to involve staff in important decisions |
| d. asks for my opinion before making decisions that affect my work | d. Senior managers act on staff feedback |
| e. is supportive in a personal crisis | |
| f. takes a positive interest in my health and well-being | |
| g. values my work | |

#### 1.3 List of non-specialist NHS hospital trusts in analysis

Table 3: List of 129 Acute NHS Trusts included in analysis presented in paper with data for 2018/19

| **Trust Name** | **Organisation Code** | **Managers (% of all staff)** | **Managers (FTE)** | **Spend on Management (£ millions)** | **Staff Survey Management Score** |
|---|---|---|---|---|---|
| Aintree University Hospital | REM | 3.27 | 148.3 | 8.89 | 63.7 |
| Airedale | RCF | 3.39 | 85 | 5.06 | 67.1 |
| Ashford and St. Peter's Hospitals | RTK | 2.44 | 79.5 | 5.39 | 64.2 |
| Barking, Havering and Redbridge University Hospitals | RF4 | 2.57 | 149.2 | 11.51 | 63.3 |
| Barnsley Hospital | RFF | 2.52 | 74.4 | 4.09 | 65.6 |
| Barts Health | R1H | 2.07 | 302.1 | 19.76 | 64.2 |
| Basildon and Thurrock University Hospitals | RDD | 3.16 | 133.3 | 8.68 | 64.0 |
| Bedford Hospital | RC1 | 2.34 | 58.5 | 3.75 | 64.0 |
| Blackpool Teaching Hospitals | RXL | 4.64 | 288.4 | 15.05 | 64.5 |
| Bolton | RMC | 1.73 | 88.5 | 5.74 | 68.4 |
| Bradford Teaching Hospitals | RAE | 2.19 | 112.8 | 7.39 | 65.3 |
| Brighton and Sussex University Hospitals | RXH | 2.63 | 192.7 | 11.47 | 64.4 |
| Buckinghamshire Healthcare | RXQ | 3.88 | 197.7 | 11.39 | 66.2 |
| Calderdale and Huddersfield | RWY | 2.42 | 125.1 | 6.69 | 63.0 |
| Cambridge University Hospitals | RGT | 2.84 | 262.8 | 16.36 | 66.6 |
| Chelsea and Westminster Hospital | RQM | 4.83 | 261.7 | 16.06 | 65.7 |
| Chesterfield Royal Hospital | RFS | 1.91 | 67.1 | 3.81 | 65.7 |
| City Hospitals Sunderland | RLN | 2.46 | 112.4 | 6.61 | 65.3 |
| Colchester Hospital University | RDE | 2.84 | 230.8 | 14.27 | 60.5 |
| Countess of Chester Hospital | RJR | 3.11 | 107.1 | 5.89 | 63.9 |
| County Durham and Darlington | RXP | 3.03 | 186.6 | 10.52 | 62.8 |
| Croydon Health Services | RJ6 | 4.31 | 139.1 | 8.15 | 62.2 |
| Dartford and Gravesham | RN7 | 4.83 | 139.4 | 7.87 | 65.5 |
| Derby Teaching Hospitals | RTG | 2.81 | 289.3 | 16.63 | 64.2 |
| Doncaster and Bassetlaw Teaching Hospitals | RP5 | 3.26 | 173.7 | 9.42 | 61.7 |

**Table 3 continued from previous page**

| **Trust Name** | **Organisation Code** | **Managers (% of all staff)** | **Managers (FTE)** | **Spend on Management (£ millions)** | **Staff Survey Management Score** |
|---|---|---|---|---|---|
| Dorset County Hospital | RBD | 3.99 | 90.1 | 5.28 | 66.1 |
| Dudley Group | RNA | 1.3 | 56.1 | 3.43 | 62.8 |
| East and North Hertfordshire | RWH | 1.45 | 72.3 | 5.32 | 61.9 |
| East Cheshire | RJN | 3.64 | 80.9 | 4.67 | 67.5 |
| East Kent Hospitals University | RVV | 1.73 | 132.8 | 8.44 | 60.5 |
| East Lancashire Hospitals | RXR | 3.24 | 238.6 | 13.36 | 67.7 |
| East Sussex Healthcare | RXC | 2.64 | 159.3 | 9.62 | 64.8 |
| Epsom and St Helier University Hospitals | RVR | 5.67 | 252.5 | 14.94 | 61.4 |
| Frimley Health | RDU | 2.21 | 177 | 11.3 | 68.3 |
| Gateshead Health | RR7 | 3.2 | 124.6 | 7.36 | 67.8 |
| George Eliot Hospital | RLT | 2.67 | 53.4 | 3.45 | 62.1 |
| Gloucestershire Hospitals | RTE | 2.16 | 140.8 | 8.45 | 62.7 |
| Great Western Hospitals | RN3 | 2.32 | 90.7 | 5.82 | 63.9 |
| Guy's and St Thomas' | RJ1 | 3.71 | 536.9 | 35.62 | 67.1 |
| Hampshire Hospitals | RN5 | 2.9 | 151 | 9.22 | 61.9 |
| Harrogate and District | RCD | 2.58 | 94.5 | 5.48 | 67.0 |
| Hillingdon Hospitals | RAS | 4.33 | 126.2 | 7.83 | 62.9 |
| Homerton University Hospital | RQX | 3.12 | 105.3 | 6.64 | 67.9 |
| Hull and East Yorkshire Hospitals | RWA | 3.6 | 259.4 | 14.42 | 65.4 |
| Imperial College Healthcare | RYJ | 4.56 | 465.5 | 27.69 | 62.6 |
| James Paget University Hospitals | RGP | 3.45 | 91.6 | 5.46 | 63.4 |
| Kettering General Hospital | RNQ | 3.64 | 125.6 | 7.1 | 65.0 |
| King's College Hospital | RJZ | 2.63 | 298.3 | 19.7 | 61.7 |
| Kingston Hospital | RAX | 3.37 | 96.8 | 6.77 | 66.9 |
| Lancashire Teaching Hospitals | RXN | 2.19 | 151.8 | 9.36 | 63.2 |
| Leeds Teaching Hospitals | RR8 | 1.57 | 237.3 | 15.77 | 68.2 |
| Lewisham and Greenwich | RJ2 | 3.56 | 197.8 | 12.36 | 65.7 |
| London North West Healthcare | R1K | 4.44 | 337.2 | 20.51 | 62.5 |

**Table 3 continued from previous page**

| Trust Name | Organisation Code | Managers (% of all staff) | Managers (FTE) | Spend on Management (£ millions) | Staff Survey Management Score |
|---|---|---|---|---|---|
| Luton and Dunstable University Hospital | RC9 | 4.25 | 158.1 | 9.33 | 67.3 |
| Maidstone and Tunbridge Wells | RWF | 3.4 | 167.2 | 9.78 | 62.8 |
| Medway | RPA | 2.43 | 89 | 5.8 | 56.5 |
| Mid Cheshire Hospitals | RBT | 2.77 | 102.8 | 5.98 | 65.4 |
| Mid Essex Hospital Services | RQ8 | 2.73 | 109.1 | 6.71 | 62.7 |
| Mid Yorkshire Hospitals | RXF | 3.35 | 237.5 | 13.19 | 63.3 |
| Milton Keynes University Hospital | RD8 | 4.51 | 133.5 | 7.99 | 64.2 |
| Newcastle Upon Tyne Hospitals | RTD | 1.34 | 170.2 | 9.08 | 66.6 |
| Norfolk and Norwich University Hospitals | RM1 | 1.97 | 133.7 | 8.46 | 61.6 |
| North Bristol | RVJ | 4 | 280.8 | 16.18 | 62.3 |
| North Cumbria University Hospitals | RNL | 3.04 | 106.9 | 6.24 | 56.8 |
| North Middlesex University Hospital | RAP | 6.1 | 181.7 | 9.92 | 64.5 |
| North Tees and Hartlepool | RVW | 3.1 | 140.2 | 8.11 | 66.4 |
| North West Anglia | RGN | 4.01 | 214.1 | 11.69 | 63.3 |
| Northampton General Hospital | RNS | 3.83 | 164 | 8.67 | 63.5 |
| Northern Devon Healthcare | RBZ | 4.38 | 111.4 | 6.48 | 68.2 |
| Northern Lincolnshire and Goole | RJL | 4.91 | 256.5 | 13.82 | 59.4 |
| Northumbria Healthcare | RTF | 4.54 | 310.1 | 17.66 | 68.0 |
| Nottingham University Hospitals | RX1 | 3.63 | 472.8 | 26.36 | 65.2 |
| Oxford University Hospitals | RTH | 2.97 | 314 | 18.56 | 63.8 |
| Pennine Acute Hospitals | RW6 | 3.37 | 295.7 | 17.12 | 62.3 |
| Plymouth Hospitals | RK9 | 2.28 | 141.8 | 8.99 | 66.4 |
| Poole Hospital | RD3 | 2.92 | 98.2 | 5.66 | 65.4 |
| Portsmouth Hospitals | RHU | 7.78 | 493.2 | 24.63 | 65.6 |
| Princess Alexandra Hospital | RQW | 5.64 | 168.3 | 10.05 | 66.4 |
| Queen Elizabeth Hospital King's Lynn | RCX | 3.96 | 110 | 6.29 | 58.7 |
| Rotherham | RFR | 3.13 | 115.1 | 6.88 | 62.2 |
| Royal Berkshire | RHW | 2.94 | 136.9 | 7.93 | 66.5 |

**Table 3 continued from previous page**

| **Trust Name** | **Organisation Code** | **Managers (% of all staff)** | **Managers (FTE)** | **Spend on Management (£ millions)** | **Staff Survey Management Score** |
|---|---|---|---|---|---|
| Royal Bournemouth and Christchurch Hospitals | RDZ | 2.98 | 118.4 | 7.23 | 69.7 |
| Royal Cornwall Hospitals | REF | 2.42 | 114 | 7.37 | 61.2 |
| Royal Devon and Exeter | RH8 | 2.42 | 167.1 | 9.61 | 66.4 |
| Royal Free London | RAL | 1.6 | 132.9 | 10.05 | 65.2 |
| Royal Liverpool and Broadgreen University Hospitals | RQ6 | 5 | 314.7 | 17.3 | 65.4 |
| Royal Surrey County Hospital | RA2 | 4.15 | 149.2 | 9.2 | 66.5 |
| Royal United Hospitals Bath | RD1 | 4 | 177.1 | 10 | 64.0 |
| Royal Wolverhampton | RL4 | 1.83 | 133 | 8.5 | 66.4 |
| Salford Royal | RM3 | 3.94 | 285 | 17.4 | 64.8 |
| Salisbury | RNZ | 2.82 | 81.2 | 4.67 | 65.1 |
| Sandwell and West Birmingham Hospitals | RXK | 3.2 | 190.2 | 11.92 | 65.6 |
| Sheffield Teaching Hospitals | RHQ | 2.26 | 315.1 | 19.57 | 65.9 |
| Sherwood Forest Hospitals | RK5 | 4.54 | 186 | 10.55 | 67.0 |
| Shrewsbury and Telford Hospital | RXW | 4.84 | 241 | 12.75 | 61.6 |
| South Tees Hospitals | RTR | 2.11 | 156.6 | 9.2 | 59.4 |
| South Tyneside | RE9 | 2.1 | 58.4 | 3.19 | 63.4 |
| South Warwickshire | RJC | 3.4 | 133.4 | 7.55 | 66.1 |
| Southend University Hospital | RAJ | 0.26 | 9.8 | 1.03 | 65.4 |
| Southport and Ormskirk Hospital | RVY | 1.9 | 47.8 | 3.33 | 59.1 |
| St George's University Hospitals | RJ7 | 2.19 | 176 | 11.94 | 61.4 |
| St Helens and Knowsley Hospitals | RBN | 3.23 | 168.2 | 12.78 | 69.7 |
| Stockport | RWJ | 3.61 | 162.9 | 9.59 | 63.1 |
| Surrey and Sussex Healthcare | RTP | 2.81 | 102.7 | 6.85 | 70.8 |
| Tameside and Glossop Integrated Care | RMP | 2.82 | 95.2 | 6.02 | 64.6 |
| Taunton and Somerset | RBA | 2.57 | 102.7 | 6.11 | 66.0 |
| Torbay and South Devon | RA9 | 3.21 | 163.4 | 9.63 | 64.8 |
| United Lincolnshire Hospitals | RWD | 2.26 | 144.8 | 8.84 | 58.5 |
| University College London Hospitals | RRV | 6.2 | 493 | 30.31 | 65.4 |

**Table 3 continued from previous page**

| Trust Name | Organisation Code | Managers (% of all staff) | Managers (FTE) | Spend on Management (£ millions) | Staff Survey Management Score |
|---|---|---|---|---|---|
| University Hospital Southampton | RHM | 5.84 | 563.3 | 28.49 | 67.9 |
| University Hospitals Birmingham | RRK | 1.85 | 327.9 | 20.13 | 63.3 |
| University Hospitals Bristol | RA7 | 2.35 | 187.7 | 11.98 | 65.4 |
| University Hospitals Coventry and Warwickshire | RKB | 4.54 | 322.7 | 17.82 | 65.9 |
| University Hospitals of Leicester | RWE | 1.5 | 193.3 | 12.79 | 63.0 |
| University Hospitals of Morecambe Bay | RTX | 3.39 | 178.6 | 10.23 | 65.4 |
| University Hospitals of North Midlands | RJE | 2.36 | 220.5 | 12.97 | 61.7 |
| Walsall Healthcare | RBK | 2.78 | 101.2 | 5.67 | 63.5 |
| Warrington and Halton Hospitals | RWW | 5.16 | 180.3 | 9.96 | 66.2 |
| West Hertfordshire Hospitals | RWG | 4.02 | 170.6 | 10.08 | 67.4 |
| West Suffolk | RGR | 4.75 | 156 | 8.55 | 67.5 |
| Western Sussex Hospitals | RYR | 2.04 | 118.1 | 8.18 | 66.1 |
| Weston Area Health | RA3 | 1.06 | 15.2 | 1.03 | 59.2 |
| Whittington Hospital | RKE | 4.43 | 162.8 | 9.68 | 64.5 |
| Wirral University Teaching Hospital | RBL | 3.39 | 176.3 | 9.63 | 60.8 |
| Worcestershire Acute Hospitals | RWP | 2.45 | 125.1 | 7.91 | 60.0 |
| Wrightington, Wigan and Leigh | RRF | 3.68 | 159.7 | 9.35 | 64.0 |
| Wye Valley | RLQ | 4.2 | 110.7 | 6.07 | 62.2 |
| Yeovil District Hospital | RA4 | 4.04 | 79.1 | 4.69 | 68.3 |
| York Teaching Hospital | RCB | 3.14 | 220.1 | 12.4 | 64.4 |

### 2 Descriptive statistics

#### 2.1 Change in number of managers over time in NHS

Figure 1: Managers time series in acute NHS hospital trusts using basic management definition ($M_1$)

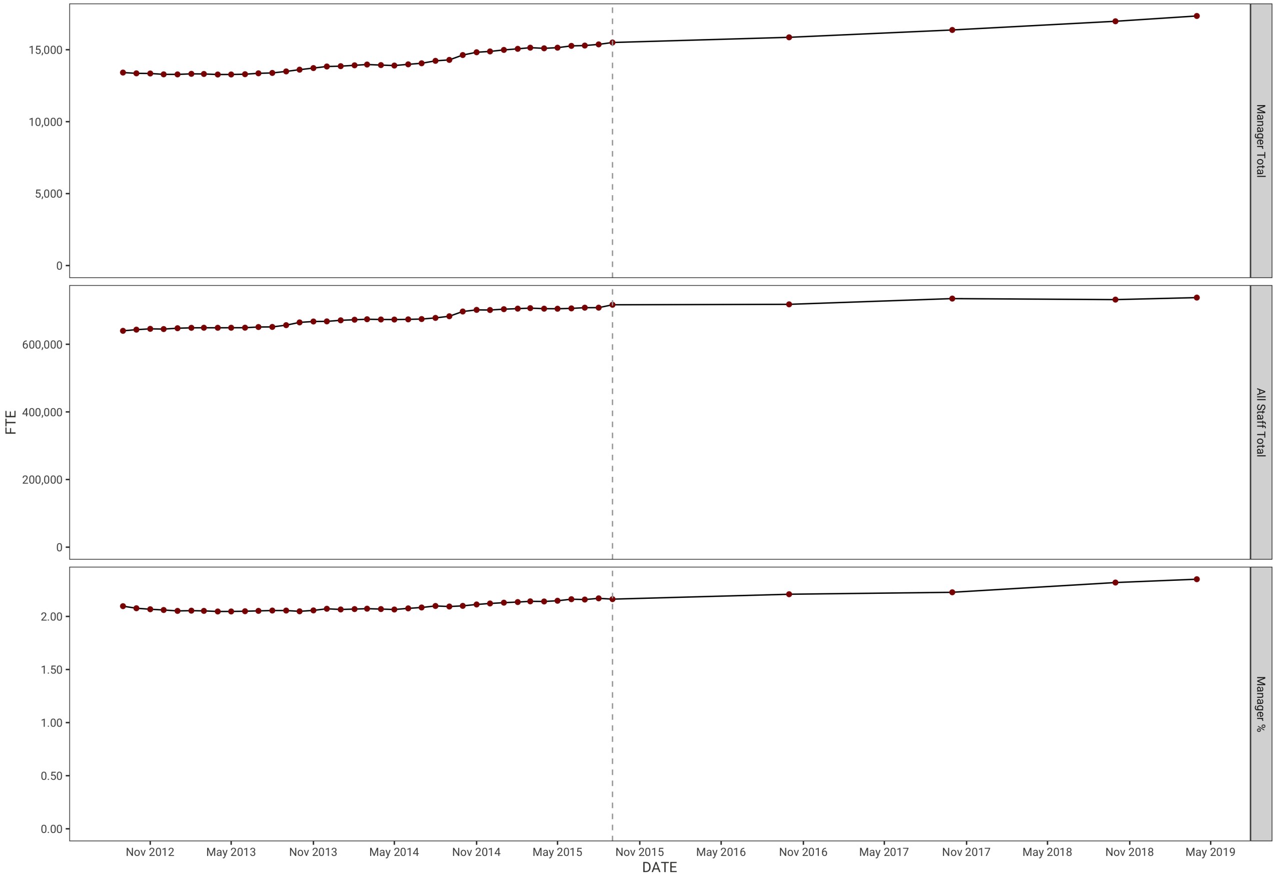

#### 2.2 Distribution of management, control and outcome variables

Figure 2: Distribution of key variables across 129 acute NHS hospital trusts in 2016/17

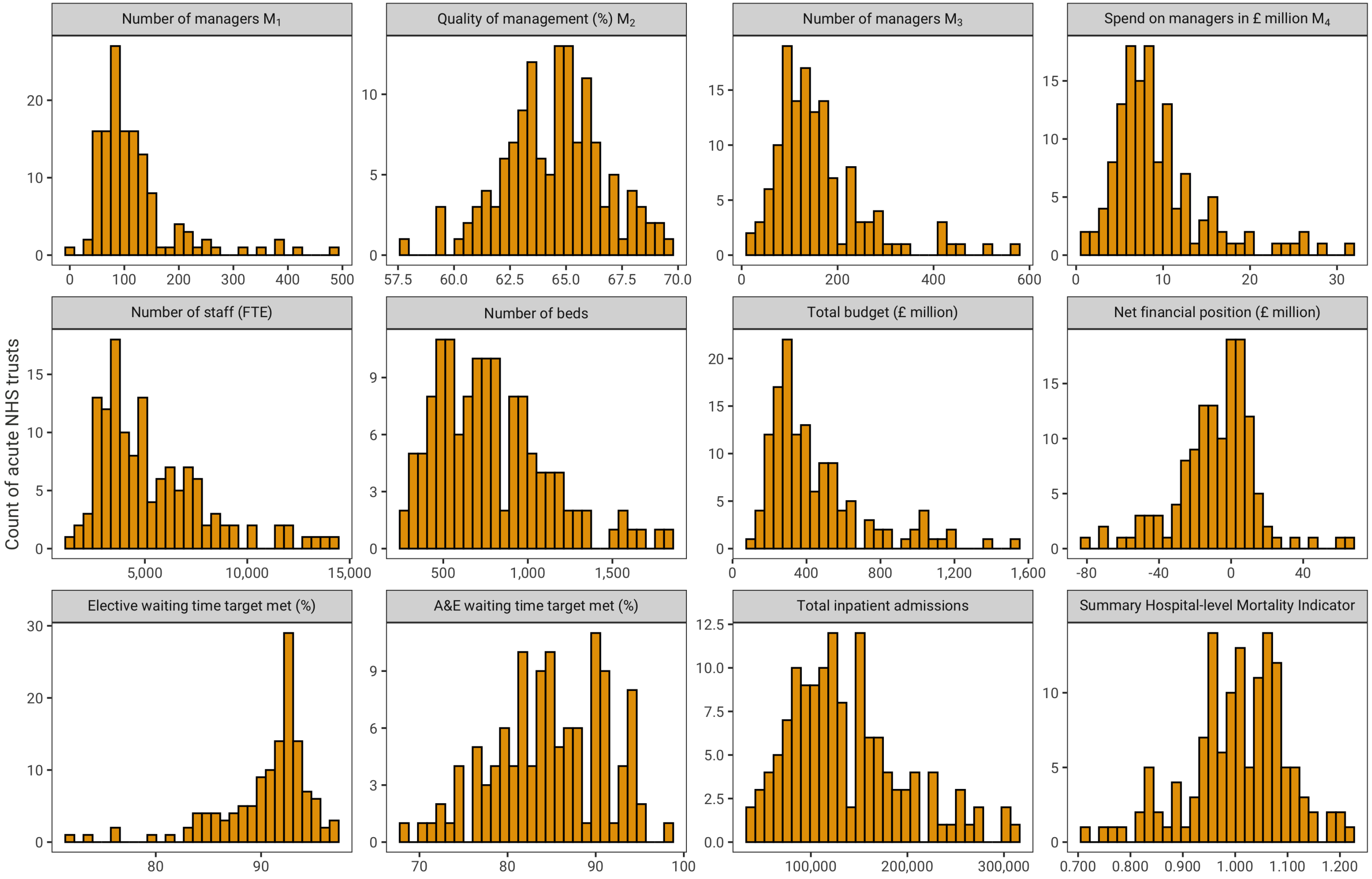

Figure 3: Distribution of key variables across 129 acute NHS hospital trusts in 2017/18

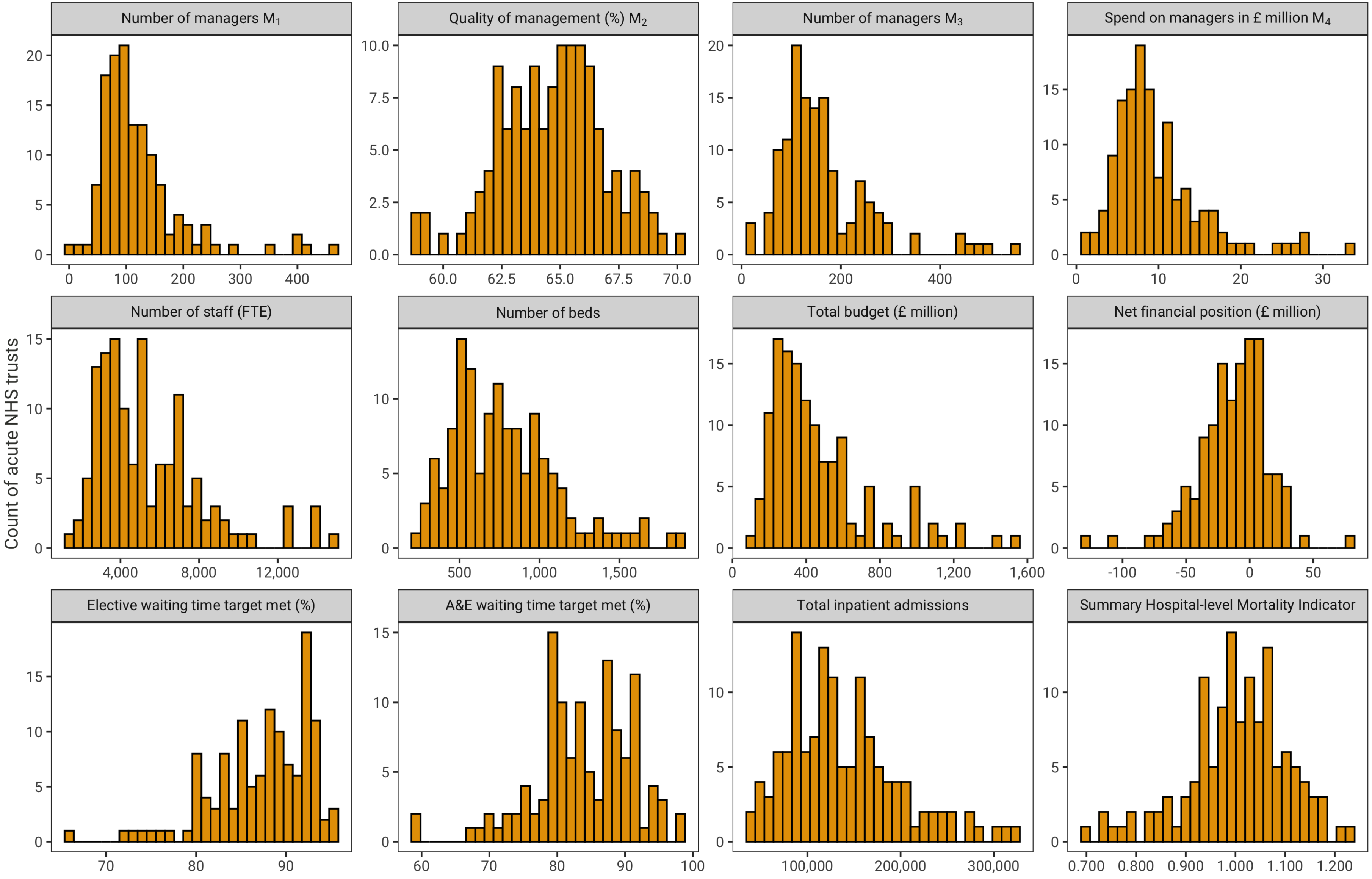

Figure 4: Distribution of key variables across 129 acute NHS hospital trusts in 2018/19

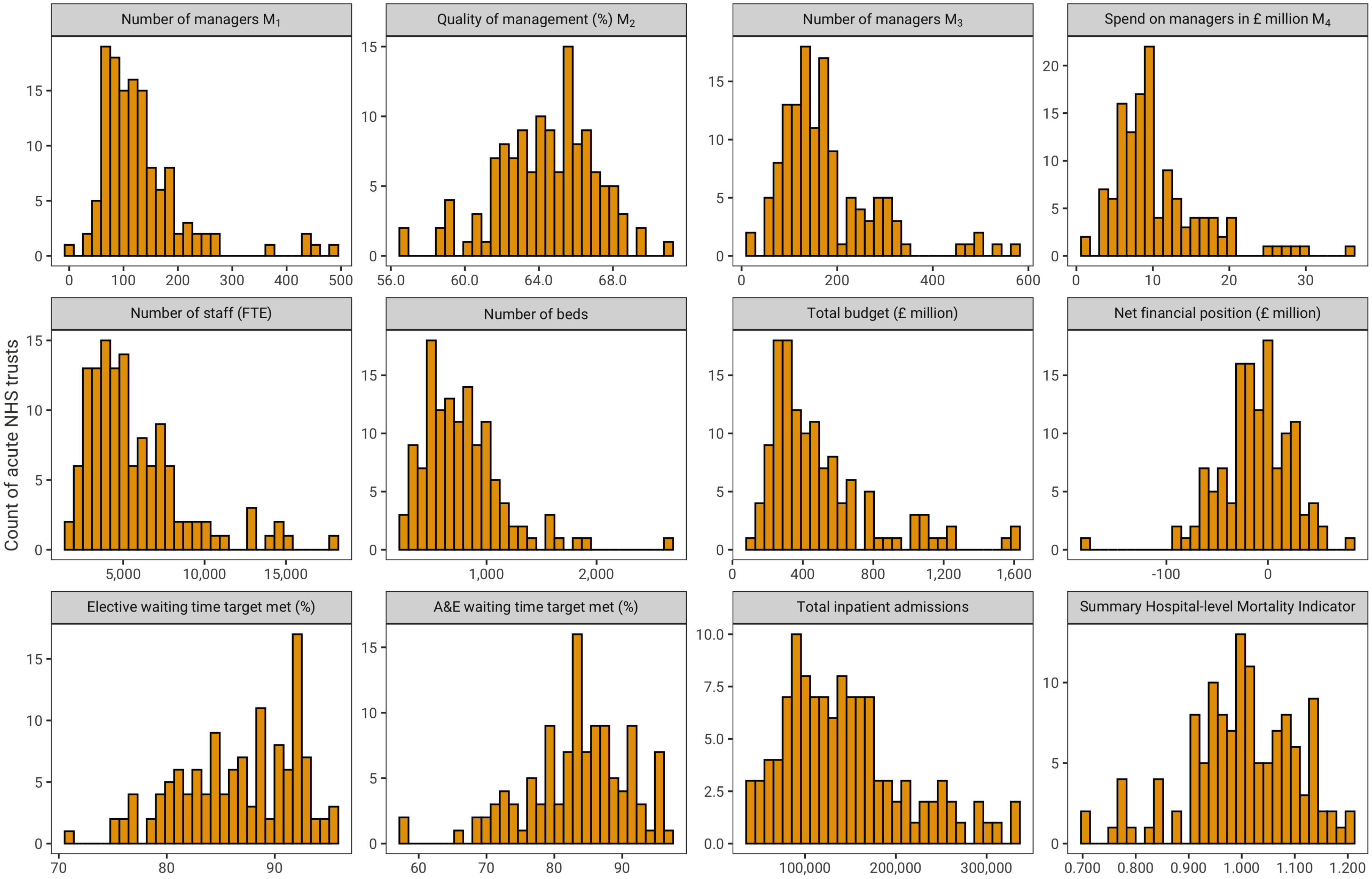

#### 2.3 Mediators between management input and outcomes

Figure 5: Relationship between management input (%) and resources to be managed across 129 acute NHS hospital trusts in 2012/13

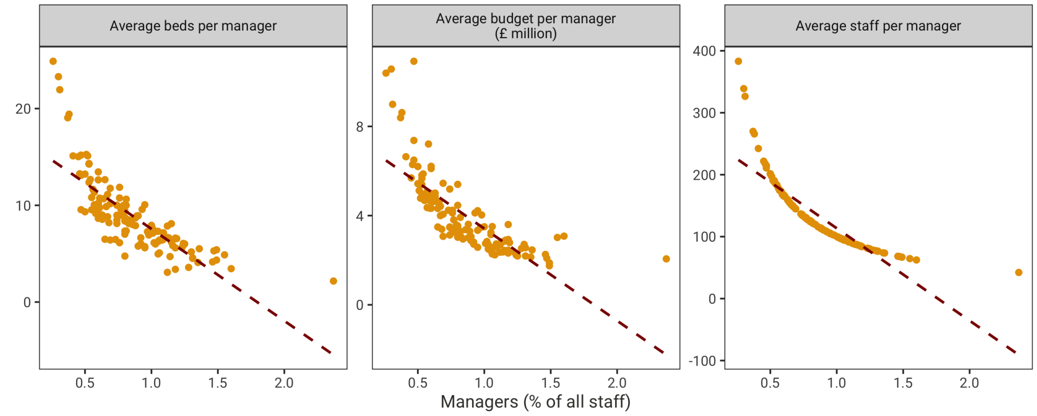

Figure 6: Relationship between management input (%) and resources to be managed across 129 acute NHS hospital trusts in 2013/14

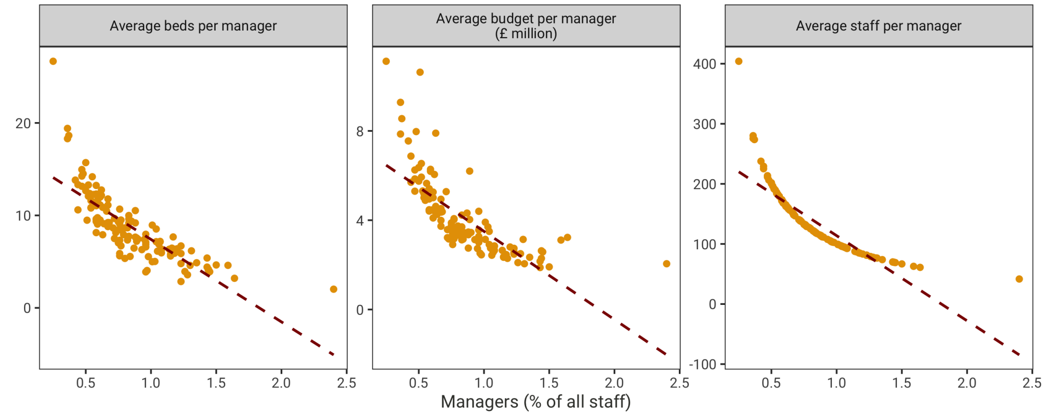

Figure 7: Relationship between management input (%) and resources to be managed across 129 acute NHS hospital trusts in 2014/15

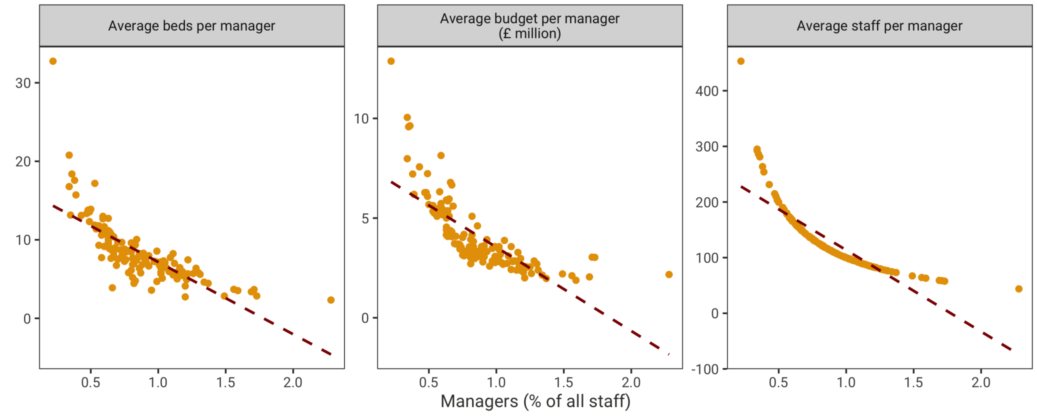

Figure 8: Relationship between management input (%) and resources to be managed across 129 acute NHS hospital trusts in 2015/16

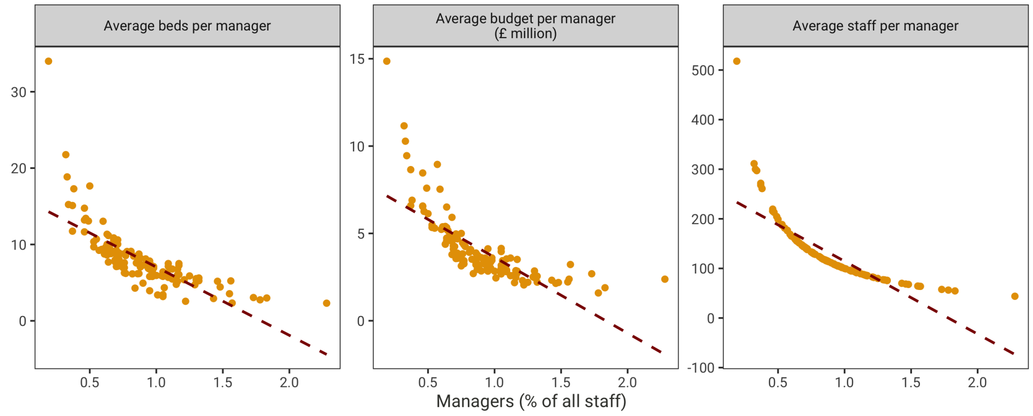

Figure 9: Relationship between management input (%) and resources to be managed across 129 acute NHS hospital trusts in 2016/17

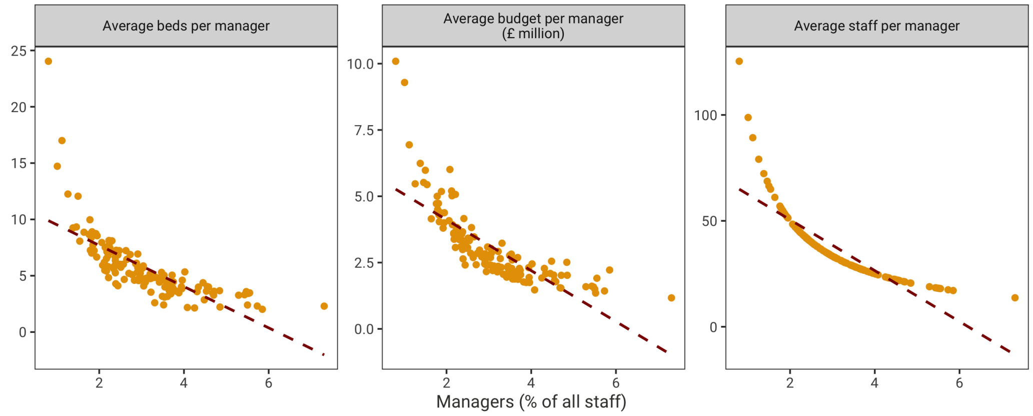

Figure 10: Relationship between management input (%) and resources to be managed across 129 acute NHS hospital trusts in 2017/18

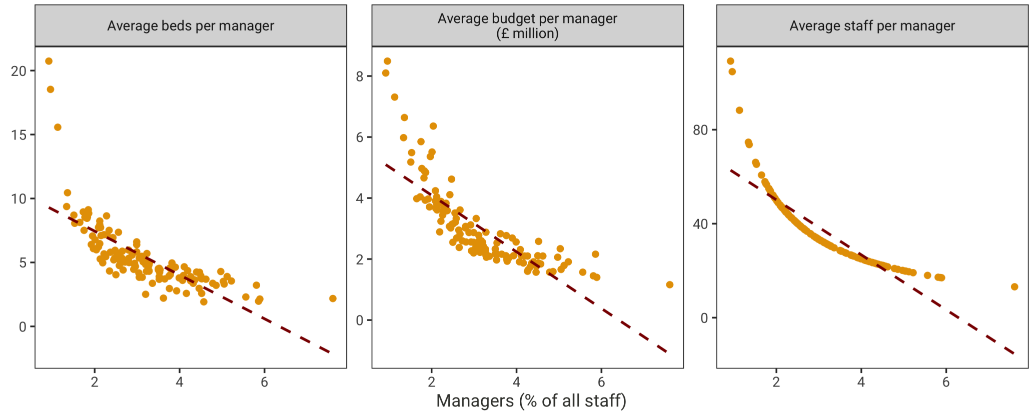

Figure 11: Relationship between management input (%) and resources to be managed across 129 acute NHS hospital trusts in 2018/19

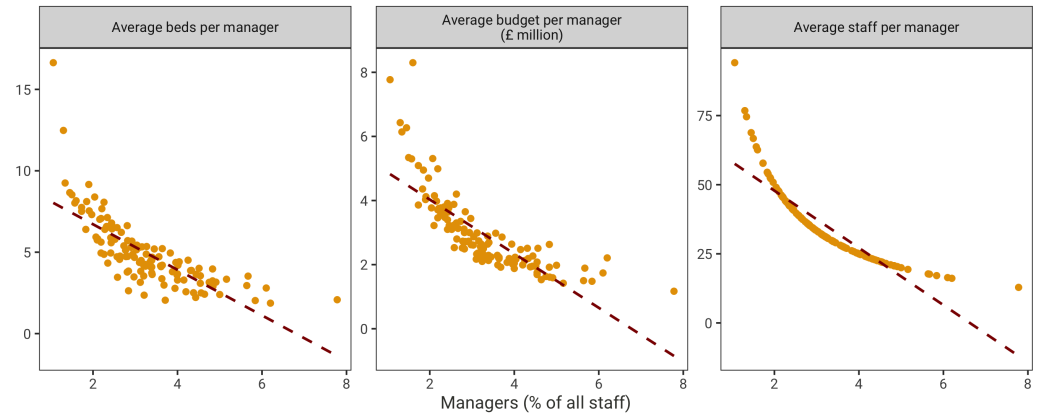

### 3 Examining between and within variation in data

#### 3.1 Comparison of variation in management measures, controls and outcomes between hospitals versus within hospitals over time

Table 4: Overall, between and within variation in management, control and outcome variables 2012/13 – 2018/19

| Variable | | Mean | Std. Dev. | Min | Max | Observations |
|---|---|---|---|---|---|---|
| Number of managers ($M_1$) | overall | 113.80 | 73.44 | 4 | 492.85 | N = 928 |
| | between | | 71.48 | 7.29 | 407.60 | n = 134 |
| | within | | 21.35 | -33.27 | 234.99 | T-bar = 6.93 |
| Quality of management ($M_2$) | overall | 62.68 | 3.21 | 48.80 | 71 | N = 927 |
| | between | | 2.08 | 55.27 | 67.59 | n = 134 |
| | within | | 2.44 | 53.23 | 68.63 | T-bar = 6.92 |
| Number of managers ($M_3$) | overall | 167.08 | 98.47 | 9.80 | 563.3 | N = 395 |
| | between | | 100.72 | 10.27 | 562.2 | n = 134 |
| | within | | 15.76 | 102.91 | 289.21 | T-bar = 2.95 |
| Spend on managers in £ million ($M_4$) | overall | 9.93 | 5.64 | 1.01 | 35.62 | N = 395 |
| | between | | 5.75 | 1.11 | 32.45 | n = 134 |
| | within | | 0.92 | 6.02 | 16.92 | T-bar = 2.95 |
| Number of staff (FTE) | overall | 9397.27 | 5939.77 | 1424 | 35251.25 | N = 923 |
| | between | | 4604.45 | 2659.53 | 25306.28 | n = 134 |
| | within | | 3821.12 | -3648.72 | 19342.24 | T-bar = 6.89 |
| Number of beds | overall | 776.98 | 344.79 | 224 | 2658 | N = 926 |
| | between | | 335.46 | 248.71 | 1894.14 | n = 134 |
| | within | | 86.66 | 313.13 | 2163.84 | T-bar = 6.91 |
| Total budget (£ million) | overall | 427.46 | 261.87 | 93.53 | 1621.49 | N = 927 |
| | between | | 255.36 | 106.46 | 1473.48 | n = 134 |
| | within | | 64.27 | 37.98 | 1196.07 | T-bar = 6.92 |
| Net financial position (£ million) | overall | -8.44 | 22.74 | -180.07 | 80.51 | N = 927 |
| | between | | 16.60 | -73.56 | 33.36 | n = 134 |
| | within | | 15.74 | -120.87 | 65.53 | T-bar = 6.92 |
| Elective waiting time target met (%) | overall | 91.15 | 4.91 | 66.20 | 99.70 | N = 896 |
| | between | | 2.79 | 83.69 | 96.40 | n = 134 |
| | within | | 4.05 | 71.15 | 105.08 | T-bar = 6.69 |
| A&E waiting time target met (%) | overall | 88.19 | 7.19 | 52 | 100 | N = 823 |
| | between | | 4.68 | 71 | 97.86 | n = 119 |
| | within | | 5.54 | 56.62 | 108.19 | T-bar = 6.92 |
| Total inpatient admissions | overall | 130909 | 58086 | 36094 | 332520 | N = 899 |
| | between | | 56190 | 38731 | 300846 | n = 134 |
| | within | | 15352 | 37776 | 247145 | T-bar = 6.71 |
| Summary Hospital-level Mortality Indicator | overall | 1.00 | 0.10 | 0.54 | 1.23 | N = 922 |
| | between | | 0.09 | 0.68 | 1.160 | n = 134 |
| | within | | 0.04 | 0.77 | 1.19 | T-bar = 6.88 |

#### 3.2 Exploring the between and within variation in the relationship between management and outcomes

Figure 12: Between variation across 129 acute NHS hospital trusts

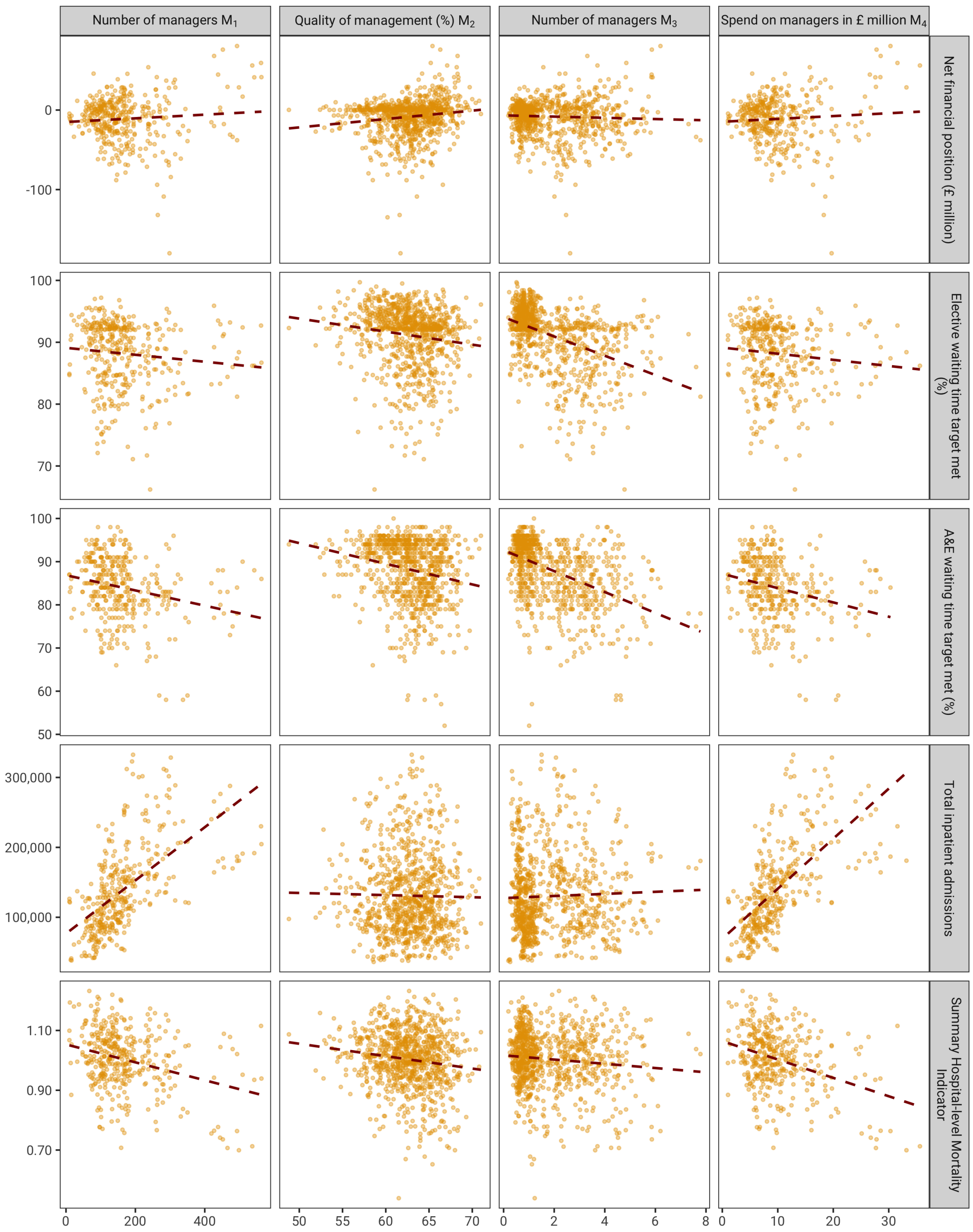

Figure 13: Within variation across 129 acute NHS hospital trusts

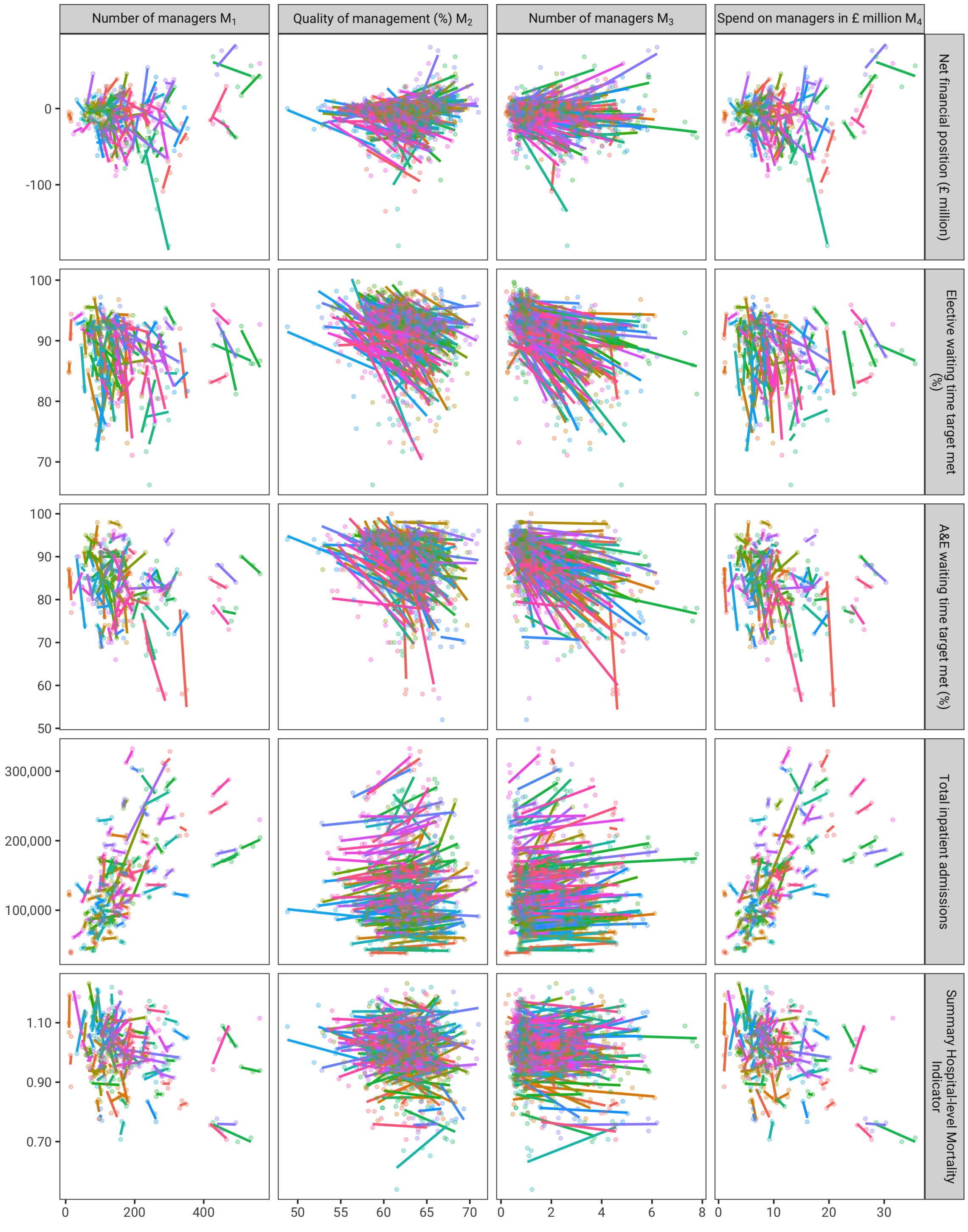

### 4 Alternative specifications of mediation regressions

#### 4.1 Number of managers ($M_1$)

Table 5: Amount of managerial input and quality of management ($M_2$) 2012/13 – 2018/19

| | Quality of management ($M_2$) | | | | | | | | | |
|---|---|---|---|---|---|---|---|---|---|---|
| | Pooled | Pooled | Between | Between | Within | Within | RE | RE | Lagged DV | Lagged DV |
| | (1) | (2) | (3) | (4) | (5) | (6) | (7) | (8) | (9) | (10) |
| Number of managers ($M_1$) | 0.003 | 0.006** | 0.003 | 0.006* | −0.005 | −0.002 | −0.0005 | 0.002 | 0.001 | 0.002* |
| | (0.002) | (0.003) | (0.003) | (0.004) | (0.004) | (0.006) | (0.002) | (0.004) | (0.001) | (0.001) |
| Size and casemix controls | FALSE | TRUE | FALSE | TRUE | FALSE | TRUE | FALSE | TRUE | FALSE | TRUE |
| Observations | 927 | 893 | 134 | 134 | 927 | 893 | 927 | 893 | 792 | 760 |
| $R^2$ | 0.356 | 0.432 | 0.013 | 0.216 | 0.004 | 0.044 | 0.586 | 0.651 | 0.702 | 0.698 |

*Note:* *p<0.1; **p<0.05; ***p<0.01
Control for number of beds, total budget, number of staff and proportion of admissions by age-sex group

Breusch-Pagan Test - RE vs pooled without controls

p = 0 therefore: RE is preferred to Pooled

Hausman Test - FE vs RE without controls

p = 0.02582 therefore: FE is preferred to RE

Breusch-Pagan Test - RE vs pooled with controls

p = 0 therefore: RE is preferred to Pooled

Hausman Test - FE vs RE with controls

p = 0.08585 therefore: RE is preferred to FE

#### 4.2 Number of managers ($M_3$)

Table 6: Amount of managerial input and quality of management ($M_2$) 2016/17 – 2018/19

| | Quality of management ($M_2$) | | | | | | | | | |
|---|---|---|---|---|---|---|---|---|---|---|
| | Pooled | Pooled | Between | Between | Within | Within | RE | RE | Lagged DV | Lagged DV |
| | (1) | (2) | (3) | (4) | (5) | (6) | (7) | (8) | (9) | (10) |
| Number of managers ($M_3$) | 0.003 | 0.005* | 0.003 | 0.005* | −0.002 | 0.006 | 0.002 | 0.004 | 0.001 | −0.0001 |
| | (0.002) | (0.003) | (0.002) | (0.003) | (0.004) | (0.005) | (0.002) | (0.002) | (0.001) | (0.001) |
| Size and casemix controls | FALSE | TRUE | FALSE | TRUE | FALSE | TRUE | FALSE | TRUE | FALSE | TRUE |
| Observations | 395 | 382 | 134 | 134 | 395 | 382 | 395 | 382 | 394 | 381 |
| $R^2$ | 0.014 | 0.141 | 0.015 | 0.177 | 0.001 | 0.107 | 0.401 | 0.552 | 0.630 | 0.635 |

*Note:* *p<0.1; **p<0.05; ***p<0.01

Control for number of beds, total budget, number of staff and proportion of admissions by age-sex group

Breusch-Pagan Test - RE vs pooled without controls

p = 0 therefore: RE is preferred to Pooled

Hausman Test - FE vs RE without controls

p = 0 therefore: FE is preferred to RE

Breusch-Pagan Test - RE vs pooled with controls

p = 0 therefore: RE is preferred to Pooled

Hausman Test - FE vs RE with controls

p = 0 therefore: FE is preferred to RE

#### 4.3 Spend on managers in £ million ($M_4$)

Table 7: Amount of managerial input and quality of management ($M_2$) 2016/17 – 2018/19

| | Quality of management ($M_2$) | | | | | | | | | |
|---|---|---|---|---|---|---|---|---|---|---|
| | Pooled | Pooled | Between | Between | Within | Within | RE | RE | Lagged DV | Lagged DV |
| | (1) | (2) | (3) | (4) | (5) | (6) | (7) | (8) | (9) | (10) |
| Spend on managers in £ million ($M_4$) | 0.046 | 0.102** | 0.047 | 0.097* | −0.066 | 0.045 | 0.026 | 0.062 | 0.012 | 0.0002 |
| | (0.032) | (0.052) | (0.033) | (0.054) | (0.071) | (0.087) | (0.030) | (0.046) | (0.012) | (0.021) |
| Size and casemix controls | FALSE | TRUE | FALSE | TRUE | FALSE | TRUE | FALSE | TRUE | FALSE | TRUE |
| Observations | 395 | 382 | 134 | 134 | 395 | 382 | 395 | 382 | 394 | 381 |
| $R^2$ | 0.014 | 0.143 | 0.015 | 0.180 | 0.003 | 0.104 | 0.398 | 0.554 | 0.630 | 0.635 |

*Note:* *p<0.1; **p<0.05; ***p<0.01
Control for number of beds, total budget, number of staff and proportion of admissions by age-sex group

Breusch-Pagan Test - RE vs pooled without controls

p = 0 therefore: RE is preferred to Pooled

Hausman Test - FE vs RE without controls

p = 0 therefore: FE is preferred to RE

Breusch-Pagan Test - RE vs pooled with controls

p = 0 therefore: RE is preferred to Pooled

Hausman Test - FE vs RE with controls

p = 0 therefore: FE is preferred to RE

### 5 Mediation of effect of management quantity on outcomes through NHS staff survey management score

#### 5.1 Number of managers ($M_1$)

Table 8: Managerial quality and Number of managers ($M_1$) 2012/13 – 2018/19

| | Quality of management ($M_2$) | | | | | | | | | | | |
|---|---|---|---|---|---|---|---|---|---|---|---|---|
| | overall | 7a | 7b | 7c | 7d | 7e | 7f | 7g | 8a | 8b | 8c | 8d |
| | (1) | (2) | (3) | (4) | (5) | (6) | (7) | (8) | (9) | (10) | (11) | (12) |
| Number of managers ($M_1$) | −0.002 | −0.002 | −0.001 | −0.001 | 0.001 | −0.002 | 0.0002 | 0.003 | 0.0003 | −0.003 | −0.001 | −0.004 |
| | (0.006) | (0.005) | (0.005) | (0.005) | (0.006) | (0.004) | (0.005) | (0.004) | (0.006) | (0.008) | (0.008) | (0.008) |
| Observations | 893 | 893 | 893 | 893 | 893 | 893 | 513 | 513 | 893 | 893 | 893 | 893 |
| $R^2$ | 0.044 | 0.028 | 0.034 | 0.028 | 0.022 | 0.021 | 0.044 | 0.032 | 0.047 | 0.042 | 0.043 | 0.044 |

*Note:* $^{*}p<0.1$; $^{**}p<0.05$; $^{***}p<0.01$
Control for hospital fixed effects, number of beds, total budget, number of staff and proportion of admissions by age-sex group

#### 5.2 Number of managers ($M_3$)

Table 9: Managerial quality and Number of managers ($M_3$) 2016/17 – 2018/19

| | Quality of management ($M_2$) | | | | | | | | | | | |
|---|---|---|---|---|---|---|---|---|---|---|---|---|
| | overall | 7a | 7b | 7c | 7d | 7e | 7f | 7g | 8a | 8b | 8c | 8d |
| | (1) | (2) | (3) | (4) | (5) | (6) | (7) | (8) | (9) | (10) | (11) | (12) |
| Number of managers ($M_3$) | 0.006 | 0.006 | 0.003 | 0.011* | 0.005 | 0.004 | −0.004 | 0.003 | 0.013* | 0.008 | 0.010 | 0.008 |
| | (0.005) | (0.006) | (0.006) | (0.006) | (0.005) | (0.005) | (0.006) | (0.006) | (0.007) | (0.009) | (0.009) | (0.010) |
| Observations | 382 | 382 | 382 | 382 | 382 | 382 | 382 | 382 | 382 | 382 | 382 | 382 |
| $R^2$ | 0.107 | 0.073 | 0.088 | 0.118 | 0.102 | 0.104 | 0.081 | 0.080 | 0.096 | 0.078 | 0.070 | 0.091 |

*Note:* $^{*}p<0.1$; $^{**}p<0.05$; $^{***}p<0.01$
Control for hospital fixed effects, number of beds, total budget, number of staff and proportion of admissions by age-sex group

#### 5.3 Spend on managers in £ million ($M_4$)

Table 10: Managerial quality and Spend on managers in £ million ($M_4$) 2016/17 – 2018/19

| | Quality of management ($M_2$) | | | | | | | | | | | |
|---|---|---|---|---|---|---|---|---|---|---|---|---|
| | overall | 7a | 7b | 7c | 7d | 7e | 7f | 7g | 8a | 8b | 8c | 8d |
| | (1) | (2) | (3) | (4) | (5) | (6) | (7) | (8) | (9) | (10) | (11) | (12) |
| Spend on managers in £ million ($M_4$) | 0.045 | 0.061 | −0.008 | 0.109 | 0.052 | 0.001 | −0.118 | −0.028 | 0.190 | 0.046 | 0.087 | 0.082 |
| | (0.087) | (0.103) | (0.093) | (0.109) | (0.083) | (0.083) | (0.117) | (0.094) | (0.116) | (0.160) | (0.149) | (0.152) |
| Observations | 382 | 382 | 382 | 382 | 382 | 382 | 382 | 382 | 382 | 382 | 382 | 382 |
| $R^2$ | 0.104 | 0.071 | 0.087 | 0.111 | 0.100 | 0.102 | 0.083 | 0.079 | 0.093 | 0.076 | 0.068 | 0.090 |

*Note:* $^{*}p<0.1$; $^{**}p<0.05$; $^{***}p<0.01$
Control for hospital fixed effects, number of beds, total budget, number of staff and proportion of admissions by age-sex group

### 6 Alternative specifications of management regressions

#### 6.1 Net financial position (£ million)

##### 6.1.1 Number of managers ($M_1$)

Table 11: Amount of managerial input and Net financial position (£ million) 2012/13 – 2018/19

| | Net financial position (£ million) | | | | | | | | | |
|---|---|---|---|---|---|---|---|---|---|---|
| | Pooled | Pooled | Between | Between | Within | Within | RE | RE | Lagged DV | Lagged DV |
| | (1) | (2) | (3) | (4) | (5) | (6) | (7) | (8) | (9) | (10) |
| Number of managers ($M_1$) | 0.020 | 0.053* | 0.021 | 0.046* | −0.007 | −0.011 | 0.012 | 0.032 | 0.004 | 0.017 |
| | (0.027) | (0.030) | (0.020) | (0.026) | (0.057) | (0.047) | (0.026) | (0.033) | (0.010) | (0.015) |
| Size and casemix controls | FALSE | TRUE | FALSE | TRUE | FALSE | TRUE | FALSE | TRUE | FALSE | TRUE |
| Observations | 927 | 893 | 134 | 134 | 927 | 893 | 927 | 893 | 792 | 760 |
| $R^2$ | 0.086 | 0.166 | 0.008 | 0.360 | 0.0001 | 0.032 | 0.145 | 0.173 | 0.614 | 0.616 |

*Note:* *p<0.1; **p<0.05; ***p<0.01
Control for number of beds, total budget, number of staff and proportion of admissions by age-sex group

Breusch-Pagan Test - RE vs pooled without controls

p = 0 therefore: RE is preferred to Pooled

Hausman Test - FE vs RE without controls

p = 0.39186 therefore: RE is preferred to FE

Breusch-Pagan Test - RE vs pooled with controls

p = 0 therefore: RE is preferred to Pooled

Hausman Test - FE vs RE with controls

p = 0.0053 therefore: FE is preferred to RE

##### 6.1.2 Quality of management ($M_2$)

Table 12: Amount of managerial input and Net financial position (£ million) 2012/13 – 2018/19

| | Net financial position (£ million) | | | | | | | | | |
|---|---|---|---|---|---|---|---|---|---|---|
| | Pooled | Pooled | Between | Between | Within | Within | RE | RE | Lagged DV | Lagged DV |
| | (1) | (2) | (3) | (4) | (5) | (6) | (7) | (8) | (9) | (10) |
| Quality of management ($M_2$) | 3.121*** | 2.844*** | 4.268*** | 3.751*** | 0.835 | 0.433 | 1.593*** | 1.209*** | 1.255*** | 1.161*** |
| | (0.364) | (0.360) | (0.585) | (0.565) | (0.561) | (0.441) | (0.433) | (0.409) | (0.214) | (0.214) |
| Size and casemix controls | FALSE | TRUE | FALSE | TRUE | FALSE | TRUE | FALSE | TRUE | FALSE | TRUE |
| Observations | 927 | 893 | 134 | 134 | 927 | 893 | 927 | 893 | 792 | 760 |
| $R^2$ | 0.208 | 0.251 | 0.287 | 0.519 | 0.008 | 0.034 | 0.169 | 0.186 | 0.629 | 0.627 |

*Note:* *p<0.1; **p<0.05; ***p<0.01
Control for number of beds, total budget, number of staff and proportion of admissions by age-sex group

Breusch-Pagan Test - RE vs pooled without controls

p = 0 therefore: RE is preferred to Pooled

Hausman Test - FE vs RE without controls

p = 0 therefore: FE is preferred to RE

Breusch-Pagan Test - RE vs pooled with controls

p = 0 therefore: RE is preferred to Pooled

Hausman Test - FE vs RE with controls

p = 3e-05 therefore: FE is preferred to RE

##### 6.1.3 Number of managers ($M_3$)

Table 13: Amount of managerial input and Net financial position (£ million) 2016/17 – 2018/19

| | Net financial position (£ million) | | | | | | | | | |
|---|---|---|---|---|---|---|---|---|---|---|
| | Pooled | Pooled | Between | Between | Within | Within | RE | RE | Lagged DV | Lagged DV |
| | (1) | (2) | (3) | (4) | (5) | (6) | (7) | (8) | (9) | (10) |
| Number of managers ($M_3$) | 0.024 | 0.062* | 0.037 | 0.066** | −0.054 | 0.124 | 0.020 | 0.076* | 0.023* | 0.023 |
| | (0.032) | (0.037) | (0.023) | (0.033) | (0.088) | (0.120) | (0.034) | (0.042) | (0.013) | (0.018) |
| Size and casemix controls | FALSE | TRUE | FALSE | TRUE | FALSE | TRUE | FALSE | TRUE | FALSE | TRUE |
| Observations | 395 | 382 | 134 | 134 | 395 | 382 | 395 | 382 | 394 | 381 |
| $R^2$ | 0.012 | 0.166 | 0.020 | 0.207 | 0.004 | 0.062 | 0.018 | 0.073 | 0.645 | 0.659 |

*Note:* *p<0.1; **p<0.05; ***p<0.01

Control for number of beds, total budget, number of staff and proportion of admissions by age-sex group

Breusch-Pagan Test - RE vs pooled without controls

p = 0 therefore: RE is preferred to Pooled

Hausman Test - FE vs RE without controls

p = 0.11259 therefore: RE is preferred to FE

Breusch-Pagan Test - RE vs pooled with controls

p = 0 therefore: RE is preferred to Pooled

Hausman Test - FE vs RE with controls

p = 0.37903 therefore: RE is preferred to FE

###### 6.1.4 Spend on managers in £ million ($M_4$)

Table 14: Amount of managerial input and Net financial position (£ million) 2016/17 – 2018/19

| | Net financial position (£ million) | | | | | | | | | |
|---|---|---|---|---|---|---|---|---|---|---|
| | Pooled | Pooled | Between | Between | Within | Within | RE | RE | Lagged DV | Lagged DV |
| | (1) | (2) | (3) | (4) | (5) | (6) | (7) | (8) | (9) | (10) |
| Spend on managers in £ million ($M_4$) | 0.376 | 1.261* | 0.599 | 1.359** | −0.916 | 2.022 | 0.312 | 1.474* | 0.413* | 0.433 |
| | (0.608) | (0.753) | (0.400) | (0.638) | (1.564) | (2.345) | (0.634) | (0.866) | (0.240) | (0.361) |
| Size and casemix controls | FALSE | TRUE | FALSE | TRUE | FALSE | TRUE | FALSE | TRUE | FALSE | TRUE |
| Observations | 395 | 382 | 134 | 134 | 395 | 382 | 395 | 382 | 394 | 381 |
| $R^2$ | 0.011 | 0.167 | 0.017 | 0.210 | 0.004 | 0.062 | 0.017 | 0.073 | 0.645 | 0.659 |

*Note:* *p<0.1; **p<0.05; ***p<0.01
Control for number of beds, total budget, number of staff and proportion of admissions by age-sex group

Breusch-Pagan Test - RE vs pooled without controls

p = 0 therefore: RE is preferred to Pooled

Hausman Test - FE vs RE without controls

p = 0.11944 therefore: RE is preferred to FE

Breusch-Pagan Test - RE vs pooled with controls

p = 0 therefore: RE is preferred to Pooled

Hausman Test - FE vs RE with controls

p = 0.32934 therefore: RE is preferred to FE

#### 6.2 Elective waiting time target met (%)

##### 6.2.1 Number of managers ($M_1$)

Table 15: Amount of managerial input and Elective waiting time target met (%) 2012/13 – 2018/19

| | Elective waiting time target met (%) | | | | | | | | | |
|---|---|---|---|---|---|---|---|---|---|---|
| | Pooled | Pooled | Between | Between | Within | Within | RE | RE | Lagged DV | Lagged DV |
| | (1) | (2) | (3) | (4) | (5) | (6) | (7) | (8) | (9) | (10) |
| Number of managers ($M_1$) | −0.005 | 0.002 | −0.006* | 0.00000 | 0.006 | 0.011 | −0.003 | 0.004 | −0.001 | 0.002 |
| | (0.003) | (0.005) | (0.003) | (0.005) | (0.006) | (0.007) | (0.004) | (0.005) | (0.002) | (0.002) |
| Size and casemix controls | FALSE | TRUE | FALSE | TRUE | FALSE | TRUE | FALSE | TRUE | FALSE | TRUE |
| Observations | 896 | 865 | 134 | 134 | 896 | 865 | 896 | 865 | 744 | 715 |
| $R^2$ | 0.361 | 0.402 | 0.022 | 0.280 | 0.002 | 0.021 | 0.543 | 0.598 | 0.635 | 0.642 |

*Note:* *p<0.1; **p<0.05; ***p<0.01

Control for number of beds, total budget, number of staff and proportion of admissions by age-sex group

Breusch-Pagan Test - RE vs pooled without controls

p = 0 therefore: RE is preferred to Pooled

Hausman Test - FE vs RE without controls

p = 0 therefore: FE is preferred to RE

Breusch-Pagan Test - RE vs pooled with controls

p = 0 therefore: RE is preferred to Pooled

Hausman Test - FE vs RE with controls

p = 0 therefore: FE is preferred to RE

##### 6.2.2 Quality of management ($M_2$)

Table 16: Amount of managerial input and Elective waiting time target met (%) 2012/13 – 2018/19

| | Elective waiting time target met (%) | | | | | | | | | |
|---|---|---|---|---|---|---|---|---|---|---|
| | Pooled | Pooled | Between | Between | Within | Within | RE | RE | Lagged DV | Lagged DV |
| | (1) | (2) | (3) | (4) | (5) | (6) | (7) | (8) | (9) | (10) |
| Quality of management ($M_2$) | 0.405*** | 0.352*** | 0.592*** | 0.547*** | 0.024 | −0.020 | 0.180** | 0.130* | 0.194*** | 0.182*** |
| | (0.071) | (0.073) | (0.104) | (0.105) | (0.097) | (0.093) | (0.078) | (0.076) | (0.045) | (0.049) |
| Size and casemix controls | FALSE | TRUE | FALSE | TRUE | FALSE | TRUE | FALSE | TRUE | FALSE | TRUE |
| Observations | 896 | 865 | 134 | 134 | 896 | 865 | 896 | 865 | 744 | 715 |
| $R^2$ | 0.400 | 0.432 | 0.196 | 0.411 | 0.0002 | 0.018 | 0.546 | 0.599 | 0.644 | 0.649 |

*Note:* *p<0.1; **p<0.05; ***p<0.01

Control for number of beds, total budget, number of staff and proportion of admissions by age-sex group

Breusch-Pagan Test - RE vs pooled without controls

p = 0 therefore: RE is preferred to Pooled

Hausman Test - FE vs RE without controls

p = 0 therefore: FE is preferred to RE

Breusch-Pagan Test - RE vs pooled with controls

p = 0 therefore: RE is preferred to Pooled

Hausman Test - FE vs RE with controls

p = 0 therefore: FE is preferred to RE

##### 6.2.3 Number of managers ($M_3$)

Table 17: Amount of managerial input and Elective waiting time target met (%) 2016/17 – 2018/19

| | Elective waiting time target met (%) | | | | | | | | | |
|---|---|---|---|---|---|---|---|---|---|---|
| | Pooled | Pooled | Between | Between | Within | Within | RE | RE | Lagged DV | Lagged DV |
| | (1) | (2) | (3) | (4) | (5) | (6) | (7) | (8) | (9) | (10) |
| Number of managers ($M_3$) | −0.005 | −0.003 | −0.004 | −0.003 | 0.003 | 0.013 | −0.003 | −0.003 | −0.003* | −0.004 |
| | (0.004) | (0.005) | (0.004) | (0.006) | (0.011) | (0.017) | (0.003) | (0.005) | (0.001) | (0.003) |
| Size and casemix controls | FALSE | TRUE | FALSE | TRUE | FALSE | TRUE | FALSE | TRUE | FALSE | TRUE |
| Observations | 384 | 371 | 134 | 134 | 384 | 371 | 384 | 371 | 373 | 360 |
| $R^2$ | 0.107 | 0.234 | 0.007 | 0.180 | 0.0004 | 0.067 | 0.515 | 0.596 | 0.613 | 0.635 |

*Note:* *p<0.1; **p<0.05; ***p<0.01

Control for number of beds, total budget, number of staff and proportion of admissions by age-sex group

Breusch-Pagan Test - RE vs pooled without controls

p = 0 therefore: RE is preferred to Pooled

Hausman Test - FE vs RE without controls

p = 0.34332 therefore: RE is preferred to FE

Breusch-Pagan Test - RE vs pooled with controls

p = 0 therefore: RE is preferred to Pooled

Hausman Test - FE vs RE with controls

p = 0.00416 therefore: FE is preferred to RE

##### 6.2.4 Spend on managers in £ million ($M_4$)

Table 18: Amount of managerial input and Elective waiting time target met (%) 2016/17 – 2018/19

| | Elective waiting time target met (%) | | | | | | | | | |
|---|---|---|---|---|---|---|---|---|---|---|
| | Pooled | Pooled | Between | Between | Within | Within | RE | RE | Lagged DV | Lagged DV |
| | (1) | (2) | (3) | (4) | (5) | (6) | (7) | (8) | (9) | (10) |
| Spend on managers in £ million ($M_4$) | −0.079 | −0.077 | −0.068 | −0.064 | 0.098 | 0.317 | −0.051 | −0.043 | −0.042* | −0.083 |
| | (0.063) | (0.112) | (0.069) | (0.112) | (0.188) | (0.275) | (0.055) | (0.106) | (0.025) | (0.051) |
| Size and casemix controls | FALSE | TRUE | FALSE | TRUE | FALSE | TRUE | FALSE | TRUE | FALSE | TRUE |
| Observations | 384 | 371 | 134 | 134 | 384 | 371 | 384 | 371 | 373 | 360 |
| $R^2$ | 0.107 | 0.234 | 0.007 | 0.181 | 0.001 | 0.069 | 0.515 | 0.595 | 0.612 | 0.635 |

*Note:* *p<0.1; **p<0.05; ***p<0.01
Control for number of beds, total budget, number of staff and proportion of admissions by age-sex group

Breusch-Pagan Test - RE vs pooled without controls

p = 0 therefore: RE is preferred to Pooled

Hausman Test - FE vs RE without controls

p = 0.34516 therefore: RE is preferred to FE

Breusch-Pagan Test - RE vs pooled with controls

p = 0 therefore: RE is preferred to Pooled

Hausman Test - FE vs RE with controls

p = 0.00483 therefore: FE is preferred to RE

#### 6.3 A&E waiting time target met (%)

##### 6.3.1 Number of managers ($M_1$)

Table 19: Amount of managerial input and A&E waiting time target met (%) 2012/13 – 2018/19

| | A&E waiting time target met (%) | | | | | | | | | |
|---|---|---|---|---|---|---|---|---|---|---|
| | Pooled | Pooled | Between | Between | Within | Within | RE | RE | Lagged DV | Lagged DV |
| | (1) | (2) | (3) | (4) | (5) | (6) | (7) | (8) | (9) | (10) |
| Number of managers ($M_1$) | −0.010* | −0.006 | −0.012* | −0.006 | −0.013 | −0.016 | −0.012** | −0.010 | −0.004 | −0.002 |
| | (0.005) | (0.006) | (0.006) | (0.010) | (0.010) | (0.010) | (0.005) | (0.007) | (0.002) | (0.003) |
| Size and casemix controls | FALSE | TRUE | FALSE | TRUE | FALSE | TRUE | FALSE | TRUE | FALSE | TRUE |
| Observations | 823 | 817 | 119 | 119 | 823 | 817 | 823 | 817 | 701 | 697 |
| $R^2$ | 0.323 | 0.363 | 0.029 | 0.225 | 0.004 | 0.034 | 0.485 | 0.499 | 0.646 | 0.654 |

*Note:* *p<0.1; **p<0.05; ***p<0.01

Control for number of beds, total budget, number of staff and proportion of admissions by age-sex group

Breusch-Pagan Test - RE vs pooled without controls

p = 0 therefore: RE is preferred to Pooled

Hausman Test - FE vs RE without controls

p = 0.84398 therefore: RE is preferred to FE

Breusch-Pagan Test - RE vs pooled with controls

p = 0 therefore: RE is preferred to Pooled

Hausman Test - FE vs RE with controls

p = 0 therefore: FE is preferred to RE

##### 6.3.2 Quality of management ($M_2$)

Table 20: Amount of managerial input and A&E waiting time target met (%) 2012/13 – 2018/19

| | A&E waiting time target met (%) | | | | | | | | | |
|---|---|---|---|---|---|---|---|---|---|---|
| | Pooled | Pooled | Between | Between | Within | Within | RE | RE | Lagged DV | Lagged DV |
| | (1) | (2) | (3) | (4) | (5) | (6) | (7) | (8) | (9) | (10) |
| Quality of management ($M_2$) | 0.376** | 0.456*** | 0.500** | 0.771*** | 0.110 | 0.122 | 0.187** | 0.189** | 0.101 | 0.144** |
| | (0.166) | (0.118) | (0.208) | (0.214) | (0.102) | (0.107) | (0.093) | (0.087) | (0.074) | (0.067) |
| Size and casemix controls | FALSE | TRUE | FALSE | TRUE | FALSE | TRUE | FALSE | TRUE | FALSE | TRUE |
| Observations | 823 | 817 | 119 | 119 | 823 | 817 | 823 | 817 | 701 | 697 |
| $R^2$ | 0.330 | 0.385 | 0.047 | 0.307 | 0.002 | 0.033 | 0.485 | 0.500 | 0.646 | 0.656 |

*Note:* *p<0.1; **p<0.05; ***p<0.01
Control for number of beds, total budget, number of staff and proportion of admissions by age-sex group

Breusch-Pagan Test - RE vs pooled without controls

p = 0 therefore: RE is preferred to Pooled

Hausman Test - FE vs RE without controls

p = 0.05094 therefore: RE is preferred to FE

Breusch-Pagan Test - RE vs pooled with controls

p = 0 therefore: RE is preferred to Pooled

Hausman Test - FE vs RE with controls

p = 0 therefore: FE is preferred to RE

##### 6.3.3 Number of managers ($M_3$)

Table 21: Amount of managerial input and A&E waiting time target met (%) 2016/17 – 2018/19

| | A&E waiting time target met (%) | | | | | | | | | |
|---|---|---|---|---|---|---|---|---|---|---|
| | Pooled | Pooled | Between | Between | Within | Within | RE | RE | Lagged DV | Lagged DV |
| | (1) | (2) | (3) | (4) | (5) | (6) | (7) | (8) | (9) | (10) |
| Number of managers ($M_3$) | −0.017*** | −0.017* | −0.018*** | −0.019** | 0.004 | −0.023 | −0.014** | −0.015 | −0.004 | −0.004 |
| | (0.007) | (0.009) | (0.006) | (0.009) | (0.018) | (0.024) | (0.006) | (0.009) | (0.003) | (0.003) |
| Size and casemix controls | FALSE | TRUE | FALSE | TRUE | FALSE | TRUE | FALSE | TRUE | FALSE | TRUE |
| Observations | 353 | 353 | 119 | 119 | 353 | 353 | 353 | 353 | 352 | 352 |
| $R^2$ | 0.062 | 0.128 | 0.066 | 0.150 | 0.0003 | 0.054 | 0.147 | 0.186 | 0.678 | 0.690 |

*Note:* *p<0.1; **p<0.05; ***p<0.01
Control for number of beds, total budget, number of staff and proportion of admissions by age-sex group

Breusch-Pagan Test - RE vs pooled without controls

p = 0 therefore: RE is preferred to Pooled

Hausman Test - FE vs RE without controls

p = 0 therefore: FE is preferred to RE

Breusch-Pagan Test - RE vs pooled with controls

p = 0 therefore: RE is preferred to Pooled

Hausman Test - FE vs RE with controls

p = 0.00767 therefore: FE is preferred to RE

##### 6.3.4 Spend on managers in £ million ($M_4$)

Table 22: Amount of managerial input and A&E waiting time target met (%) 2016/17 – 2018/19

| | A&E waiting time target met (%) | | | | | | | | | |
|---|---|---|---|---|---|---|---|---|---|---|
| | Pooled | Pooled | Between | Between | Within | Within | RE | RE | Lagged DV | Lagged DV |
| | (1) | (2) | (3) | (4) | (5) | (6) | (7) | (8) | (9) | (10) |
| Spend on managers in £ million ($M_4$) | −0.323*** | −0.359** | −0.334*** | −0.399** | −0.042 | −0.604 | −0.286** | −0.352* | −0.077 | −0.093 |
| | (0.113) | (0.181) | (0.111) | (0.181) | (0.322) | (0.417) | (0.112) | (0.180) | (0.050) | (0.071) |
| Size and casemix controls | FALSE | TRUE | FALSE | TRUE | FALSE | TRUE | FALSE | TRUE | FALSE | TRUE |
| Observations | 353 | 353 | 119 | 119 | 353 | 353 | 353 | 353 | 352 | 352 |
| $R^2$ | 0.068 | 0.132 | 0.072 | 0.155 | 0.0001 | 0.062 | 0.152 | 0.188 | 0.679 | 0.691 |

*Note:* *p<0.1; **p<0.05; ***p<0.01
Control for number of beds, total budget, number of staff and proportion of admissions by age-sex group

Breusch-Pagan Test - RE vs pooled without controls

p = 0 therefore: RE is preferred to Pooled

Hausman Test - FE vs RE without controls

p = 0 therefore: FE is preferred to RE

Breusch-Pagan Test - RE vs pooled with controls

p = 0 therefore: RE is preferred to Pooled

Hausman Test - FE vs RE with controls

p = 0.00321 therefore: FE is preferred to RE

#### 6.4 Total inpatient admissions

##### 6.4.1 Number of managers ($M_1$)

Table 23: Amount of managerial input and Total inpatient admissions 2012/13 – 2018/19

| | Total inpatient admissions | | | | | | | | | |
|---|---|---|---|---|---|---|---|---|---|---|
| | Pooled | Pooled | Between | Between | Within | Within | RE | RE | Lagged DV | Lagged DV |
| | (1) | (2) | (3) | (4) | (5) | (6) | (7) | (8) | (9) | (10) |
| Number of managers ($M_1$) | 465.793*** | 7.290 | 467.450*** | 9.781 | 205.396*** | 16.981 | 243.579*** | 12.405 | 21.840*** | 4.719 |
| | (64.305) | (40.084) | (55.357) | (33.194) | (64.956) | (30.135) | (57.095) | (27.222) | (7.151) | (8.338) |
| Size and casemix controls | FALSE | TRUE | FALSE | TRUE | FALSE | TRUE | FALSE | TRUE | FALSE | TRUE |
| Observations | 899 | 893 | 134 | 134 | 899 | 893 | 899 | 893 | 749 | 745 |
| $R^2$ | 0.351 | 0.885 | 0.351 | 0.913 | 0.084 | 0.606 | 0.371 | 0.745 | 0.965 | 0.968 |

*Note:* *p<0.1; **p<0.05; ***p<0.01
Control for number of beds, total budget, number of staff and proportion of admissions by age-sex group

Breusch-Pagan Test - RE vs pooled without controls

p = 0 therefore: RE is preferred to Pooled

Hausman Test - FE vs RE without controls

p = 6e-05 therefore: FE is preferred to RE

Breusch-Pagan Test - RE vs pooled with controls

p = 0 therefore: RE is preferred to Pooled

Hausman Test - FE vs RE with controls

p = 0 therefore: FE is preferred to RE

##### 6.4.2 Quality of management ($M_2$)

Table 24: Amount of managerial input and Total inpatient admissions 2012/13 – 2018/19

| | Total inpatient admissions | | | | | | | | | |
|---|---|---|---|---|---|---|---|---|---|---|
| | Pooled | Pooled | Between | Between | Within | Within | RE | RE | Lagged DV | Lagged DV |
| | (1) | (2) | (3) | (4) | (5) | (6) | (7) | (8) | (9) | (10) |
| Quality of management ($M_2$) | −2,930.598** | −466.941 | −3,897.992* | −540.942 | −1,167.568** | −80.677 | −1,214.839** | −182.481 | 148.641 | −75.529 |
| | (1,388.259) | (433.209) | (2,318.771) | (816.263) | (533.118) | (224.766) | (524.251) | (217.796) | (181.704) | (178.410) |
| Size and casemix controls | FALSE | TRUE | FALSE | TRUE | FALSE | TRUE | FALSE | TRUE | FALSE | TRUE |
| Observations | 899 | 893 | 134 | 134 | 899 | 893 | 899 | 893 | 749 | 745 |
| $R^2$ | 0.040 | 0.885 | 0.021 | 0.913 | 0.020 | 0.605 | 0.308 | 0.746 | 0.964 | 0.968 |

*Note:* *p<0.1; **p<0.05; ***p<0.01

Control for number of beds, total budget, number of staff and proportion of admissions by age-sex group

Breusch-Pagan Test - RE vs pooled without controls

p = 0 therefore: RE is preferred to Pooled

Hausman Test - FE vs RE without controls

p = 0.21314 therefore: RE is preferred to FE

Breusch-Pagan Test - RE vs pooled with controls

p = 0 therefore: RE is preferred to Pooled

Hausman Test - FE vs RE with controls

p = 0 therefore: FE is preferred to RE

##### 6.4.3 Number of managers ($M_3$)

Table 25: Amount of managerial input and Total inpatient admissions 2016/17 – 2018/19

| | Total inpatient admissions | | | | | | | | | |
|---|---|---|---|---|---|---|---|---|---|---|
| | Pooled | Pooled | Between | Between | Within | Within | RE | RE | Lagged DV | Lagged DV |
| | (1) | (2) | (3) | (4) | (5) | (6) | (7) | (8) | (9) | (10) |
| Number of managers ($M_3$) | 379.882*** | −7.277 | 360.834*** | −12.737 | 299.485** | −46.927 | 329.424*** | −37.985 | 17.716** | 12.758* |
| | (52.668) | (29.271) | (42.826) | (25.185) | (148.081) | (40.476) | (79.560) | (32.397) | (7.058) | (7.440) |
| Size and casemix controls | FALSE | TRUE | FALSE | TRUE | FALSE | TRUE | FALSE | TRUE | FALSE | TRUE |
| Observations | 382 | 382 | 134 | 134 | 382 | 382 | 382 | 382 | 381 | 381 |
| $R^2$ | 0.358 | 0.897 | 0.350 | 0.910 | 0.177 | 0.656 | 0.345 | 0.722 | 0.972 | 0.974 |

*Note:* *p<0.1; **p<0.05; ***p<0.01
Control for number of beds, total budget, number of staff and proportion of admissions by age-sex group

Breusch-Pagan Test - RE vs pooled without controls

p = 0 therefore: RE is preferred to Pooled

Hausman Test - FE vs RE without controls

p = 0.45683 therefore: RE is preferred to FE

Breusch-Pagan Test - RE vs pooled with controls

p = 0 therefore: RE is preferred to Pooled

Hausman Test - FE vs RE with controls

p = 0 therefore: FE is preferred to RE

##### 6.4.4 Spend on managers in £ million ($M_4$)

Table 26: Amount of managerial input and Total inpatient admissions 2016/17 – 2018/19

| | Total inpatient admissions | | | | | | | | | |
|---|---|---|---|---|---|---|---|---|---|---|
| | Pooled | Pooled | Between | Between | Within | Within | RE | RE | Lagged DV | Lagged DV |
| | (1) | (2) | (3) | (4) | (5) | (6) | (7) | (8) | (9) | (10) |
| Spend on managers in £ million ($M_4$) | 7,146.755*** | −93.800 | 6,733.398*** | −224.583 | 5,521.627** | −453.417 | 6,112.432*** | −489.215 | 364.667*** | 299.041** |
| | (934.153) | (569.005) | (726.613) | (494.799) | (2,536.941) | (686.583) | (1,377.492) | (558.165) | (139.232) | (150.472) |
| Size and casemix controls | FALSE | TRUE | FALSE | TRUE | FALSE | TRUE | FALSE | TRUE | FALSE | TRUE |
| Observations | 382 | 382 | 134 | 134 | 382 | 382 | 382 | 382 | 381 | 381 |
| $R^2$ | 0.409 | 0.897 | 0.394 | 0.910 | 0.202 | 0.654 | 0.374 | 0.721 | 0.973 | 0.974 |

*Note:* *p<0.1; **p<0.05; ***p<0.01

Control for number of beds, total budget, number of staff and proportion of admissions by age-sex group

Breusch-Pagan Test - RE vs pooled without controls

p = 0 therefore: RE is preferred to Pooled

Hausman Test - FE vs RE without controls

p = 0.60647 therefore: RE is preferred to FE

Breusch-Pagan Test - RE vs pooled with controls

p = 0 therefore: RE is preferred to Pooled

Hausman Test - FE vs RE with controls

p = 0 therefore: FE is preferred to RE

#### 6.5 Summary Hospital-level Mortality Indicator

##### 6.5.1 Number of managers ($M_1$)

Table 27: Amount of managerial input and Summary Hospital-level Mortality Indicator 2012/13 – 2018/19

| | Summary Hospital-level Mortality Indicator | | | | | | | | | |
|---|---|---|---|---|---|---|---|---|---|---|
| | Pooled | Pooled | Between | Between | Within | Within | RE | RE | Lagged DV | Lagged DV |
| | (1) | (2) | (3) | (4) | (5) | (6) | (7) | (8) | (9) | (10) |
| Number of managers ($M_1$) | −0.0004*** | −0.0001 | −0.0004*** | −0.0001 | −0.00002 | 0.00000 | −0.0002 | −0.00003 | −0.0001* | −0.00003 |
| | (0.0001) | (0.0001) | (0.0001) | (0.0001) | (0.0001) | (0.0001) | (0.0001) | (0.0001) | (0.00003) | (0.00003) |
| Size and casemix controls | FALSE | TRUE | FALSE | TRUE | FALSE | TRUE | FALSE | TRUE | FALSE | TRUE |
| Observations | 922 | 888 | 134 | 134 | 922 | 888 | 922 | 888 | 787 | 755 |
| $R^2$ | 0.101 | 0.445 | 0.132 | 0.594 | 0.0001 | 0.034 | 0.026 | 0.137 | 0.739 | 0.751 |

*Note:* *p<0.1; **p<0.05; ***p<0.01

Control for number of beds, total budget, number of staff and proportion of admissions by age-sex group

Breusch-Pagan Test - RE vs pooled without controls

p = 0 therefore: RE is preferred to Pooled

Hausman Test - FE vs RE without controls

p = 0.00198 therefore: FE is preferred to RE

Breusch-Pagan Test - RE vs pooled with controls

p = 0 therefore: RE is preferred to Pooled

Hausman Test - FE vs RE with controls

p = 0.00029 therefore: FE is preferred to RE

##### 6.5.2 Quality of management ($M_2$)

Table 28: Amount of managerial input and Summary Hospital-level Mortality Indicator 2012/13 – 2018/19

| | Summary Hospital-level Mortality Indicator | | | | | | | | | |
|---|---|---|---|---|---|---|---|---|---|---|
| | Pooled | Pooled | Between | Between | Within | Within | RE | RE | Lagged DV | Lagged DV |
| | (1) | (2) | (3) | (4) | (5) | (6) | (7) | (8) | (9) | (10) |
| Quality of management ($M_2$) | $-0.006^{***}$ | $-0.001$ | $-0.010^{***}$ | $-0.001$ | $-0.0001$ | 0.0001 | $-0.001$ | $-0.0003$ | 0.0001 | 0.001 |
| | (0.002) | (0.002) | (0.003) | (0.003) | (0.001) | (0.001) | (0.001) | (0.001) | (0.001) | (0.001) |
| Size and casemix controls | FALSE | TRUE | FALSE | TRUE | FALSE | TRUE | FALSE | TRUE | FALSE | TRUE |
| Observations | 922 | 888 | 134 | 134 | 922 | 888 | 922 | 888 | 787 | 755 |
| $R^2$ | 0.030 | 0.442 | 0.058 | 0.589 | 0.00002 | 0.034 | 0.019 | 0.136 | 0.738 | 0.751 |

*Note:* $^{*}p<0.1$; $^{**}p<0.05$; $^{***}p<0.01$

Control for number of beds, total budget, number of staff and proportion of admissions by age-sex group

Breusch-Pagan Test - RE vs pooled without controls

p = 0 therefore: RE is preferred to Pooled

Hausman Test - FE vs RE without controls

p = 0.01217 therefore: FE is preferred to RE

Breusch-Pagan Test - RE vs pooled with controls

p = 0 therefore: RE is preferred to Pooled

Hausman Test - FE vs RE with controls

p = 0.00035 therefore: FE is preferred to RE

##### 6.5.3 Number of managers ($M_3$)

Table 29: Amount of managerial input and Summary Hospital-level Mortality Indicator 2016/17 – 2018/19

| | Summary Hospital-level Mortality Indicator | | | | | | | | | |
|---|---|---|---|---|---|---|---|---|---|---|
| | Pooled | Pooled | Between | Between | Within | Within | RE | RE | Lagged DV | Lagged DV |
| | (1) | (2) | (3) | (4) | (5) | (6) | (7) | (8) | (9) | (10) |
| Number of managers ($M_3$) | −0.0003*** | −0.0001 | −0.0003*** | −0.00004 | −0.0001 | −0.0003 | −0.0002** | −0.0001 | −0.00001 | 0.00001 |
| | (0.0001) | (0.0001) | (0.0001) | (0.0001) | (0.0001) | (0.0002) | (0.0001) | (0.0001) | (0.00003) | (0.00003) |
| Size and casemix controls | FALSE | TRUE | FALSE | TRUE | FALSE | TRUE | FALSE | TRUE | FALSE | TRUE |
| Observations | 390 | 377 | 134 | 134 | 390 | 377 | 390 | 377 | 389 | 376 |
| $R^2$ | 0.088 | 0.456 | 0.077 | 0.494 | 0.002 | 0.040 | 0.214 | 0.415 | 0.781 | 0.794 |

*Note:* *p<0.1; **p<0.05; ***p<0.01
Control for number of beds, total budget, number of staff and proportion of admissions by age-sex group

Breusch-Pagan Test - RE vs pooled without controls

p = 0 therefore: RE is preferred to Pooled

Hausman Test - FE vs RE without controls

p = 0 therefore: FE is preferred to RE

Breusch-Pagan Test - RE vs pooled with controls

p = 0 therefore: RE is preferred to Pooled

Hausman Test - FE vs RE with controls

p = 0.00011 therefore: FE is preferred to RE

##### 6.5.4 Spend on managers in £ million ($M_4$)

Table 30: Amount of managerial input and Summary Hospital-level Mortality Indicator 2016/17 – 2018/19

| | Summary Hospital-level Mortality Indicator | | | | | | | | | |
|---|---|---|---|---|---|---|---|---|---|---|
| | Pooled | Pooled | Between | Between | Within | Within | RE | RE | Lagged DV | Lagged DV |
| | (1) | (2) | (3) | (4) | (5) | (6) | (7) | (8) | (9) | (10) |
| Spend on managers in £ million ($M_4$) | $-0.006^{***}$ | $-0.002$ | $-0.005^{***}$ | $-0.001$ | $-0.002$ | $-0.005$ | $-0.005^{***}$ | $-0.002$ | $-0.0003$ | 0.0003 |
| | (0.002) | (0.002) | (0.001) | (0.002) | (0.002) | (0.004) | (0.002) | (0.002) | (0.0005) | (0.001) |
| Size and casemix controls | FALSE | TRUE | FALSE | TRUE | FALSE | TRUE | FALSE | TRUE | FALSE | TRUE |
| Observations | 390 | 377 | 134 | 134 | 390 | 377 | 390 | 377 | 389 | 376 |
| $R^2$ | 0.121 | 0.456 | 0.110 | 0.494 | 0.002 | 0.040 | 0.224 | 0.419 | 0.781 | 0.794 |

*Note:* $^{*}p<0.1$; $^{**}p<0.05$; $^{***}p<0.01$
Control for number of beds, total budget, number of staff and proportion of admissions by age-sex group

Breusch-Pagan Test - RE vs pooled without controls

p = 0 therefore: RE is preferred to Pooled

Hausman Test - FE vs RE without controls

p = 0 therefore: FE is preferred to RE

Breusch-Pagan Test - RE vs pooled with controls

p = 0 therefore: RE is preferred to Pooled

Hausman Test - FE vs RE with controls

p = 0.00015 therefore: FE is preferred to RE

### 7 Impact of responses to individual questions within the NHS staff survey on outcomes

#### 7.1 Net financial position (£ million)

Table 31: Managerial quality and Net financial position (£ million) 2012/13 – 2018/19

| | Net financial position (£ million) | | | | | | | | | | | |
|---|---|---|---|---|---|---|---|---|---|---|---|---|
| | overall | 7a | 7b | 7c | 7d | 7e | 7f | 7g | 8a | 8b | 8c | 8d |
| | (1) | (2) | (3) | (4) | (5) | (6) | (7) | (8) | (9) | (10) | (11) | (12) |
| overall | 0.433 | | | | | | | | | | | |
| | (0.441) | | | | | | | | | | | |
| 7a | | 0.225 | | | | | | | | | | |
| | | (0.463) | | | | | | | | | | |
| 7b | | | 0.297 | | | | | | | | | |
| | | | (0.393) | | | | | | | | | |
| 7c | | | | 0.272 | | | | | | | | |
| | | | | (0.389) | | | | | | | | |
| 7d | | | | | 0.301 | | | | | | | |
| | | | | | (0.385) | | | | | | | |
| 7e | | | | | | 0.375 | | | | | | |
| | | | | | | (0.473) | | | | | | |
| 7f | | | | | | | 1.114* | | | | | |
| | | | | | | | (0.614) | | | | | |
| 7g | | | | | | | | 1.258 | | | | |
| | | | | | | | | (0.850) | | | | |
| 8a | | | | | | | | | 0.968*** | | | |
| | | | | | | | | | (0.346) | | | |
| 8b | | | | | | | | | | 0.357 | | |
| | | | | | | | | | | (0.260) | | |
| 8c | | | | | | | | | | | 0.431 | |
| | | | | | | | | | | | (0.280) | |
| 8d | | | | | | | | | | | | 0.336 |
| | | | | | | | | | | | | (0.274) |
| Observations | 893 | 893 | 893 | 893 | 893 | 893 | 513 | 513 | 893 | 893 | 893 | 893 |
| $R^2$ | 0.034 | 0.032 | 0.033 | 0.033 | 0.033 | 0.033 | 0.039 | 0.040 | 0.047 | 0.035 | 0.037 | 0.035 |

*Note:* *p<0.1; **p<0.05; ***p<0.01

Control for hospital fixed effects, number of beds, total budget, number of staff and proportion of admissions by age-sex group

#### 7.2 Elective waiting time target met (%)

Table 32: Managerial quality and Elective waiting time target met (%) 2012/13 – 2018/19

| | Elective waiting time target met (%) | | | | | | | | | | | |
|---|---|---|---|---|---|---|---|---|---|---|---|---|
| | overall | 7a | 7b | 7c | 7d | 7e | 7f | 7g | 8a | 8b | 8c | 8d |
| | (1) | (2) | (3) | (4) | (5) | (6) | (7) | (8) | (9) | (10) | (11) | (12) |
| overall | −0.020 | | | | | | | | | | | |
| | (0.093) | | | | | | | | | | | |
| 7a | | −0.003 | | | | | | | | | | |
| | | (0.087) | | | | | | | | | | |
| 7b | | | −0.025 | | | | | | | | | |
| | | | (0.079) | | | | | | | | | |
| 7c | | | | −0.017 | | | | | | | | |
| | | | | (0.080) | | | | | | | | |
| 7d | | | | | 0.055 | | | | | | | |
| | | | | | (0.073) | | | | | | | |
| 7e | | | | | | −0.005 | | | | | | |
| | | | | | | (0.091) | | | | | | |
| 7f | | | | | | | 0.177 | | | | | |
| | | | | | | | (0.108) | | | | | |
| 7g | | | | | | | | 0.171 | | | | |
| | | | | | | | | (0.129) | | | | |
| 8a | | | | | | | | | 0.0002 | | | |
| | | | | | | | | | (0.085) | | | |
| 8b | | | | | | | | | | −0.0003 | | |
| | | | | | | | | | | (0.063) | | |
| 8c | | | | | | | | | | | 0.012 | |
| | | | | | | | | | | | (0.065) | |
| 8d | | | | | | | | | | | | −0.017 |
| | | | | | | | | | | | | (0.064) |
| Observations | 865 | 865 | 865 | 865 | 865 | 865 | 497 | 497 | 865 | 865 | 865 | 865 |
| $R^2$ | 0.018 | 0.017 | 0.018 | 0.017 | 0.018 | 0.017 | 0.064 | 0.063 | 0.017 | 0.017 | 0.017 | 0.018 |

*Note:* $^{*}$p<0.1; $^{**}$p<0.05; $^{***}$p<0.01

Control for hospital fixed effects, number of beds, total budget, number of staff and proportion of admissions by age-sex group

#### 7.3 A&E waiting time target met (%)

Table 33: Managerial quality and A&E waiting time target met (%) 2012/13 – 2018/19

| | A&E waiting time target met (%) | | | | | | | | | | | |
|---|---|---|---|---|---|---|---|---|---|---|---|---|
| | overall | 7a | 7b | 7c | 7d | 7e | 7f | 7g | 8a | 8b | 8c | 8d |
| | (1) | (2) | (3) | (4) | (5) | (6) | (7) | (8) | (9) | (10) | (11) | (12) |
| overall | 0.122<br>(0.107) | | | | | | | | | | | |
| 7a | | 0.150<br>(0.111) | | | | | | | | | | |
| 7b | | | 0.185*<br>(0.106) | | | | | | | | | |
| 7c | | | | 0.126<br>(0.105) | | | | | | | | |
| 7d | | | | | 0.078<br>(0.093) | | | | | | | |
| 7e | | | | | | 0.103<br>(0.111) | | | | | | |
| 7f | | | | | | | 0.315*<br>(0.166) | | | | | |
| 7g | | | | | | | | 0.274<br>(0.176) | | | | |
| 8a | | | | | | | | | 0.008<br>(0.080) | | | |
| 8b | | | | | | | | | | 0.105<br>(0.071) | | |
| 8c | | | | | | | | | | | 0.097<br>(0.077) | |
| 8d | | | | | | | | | | | | 0.099<br>(0.081) |
| Observations | 817 | 817 | 817 | 817 | 817 | 817 | 469 | 469 | 817 | 817 | 817 | 817 |
| $R^2$ | 0.033 | 0.033 | 0.035 | 0.033 | 0.031 | 0.031 | 0.041 | 0.037 | 0.030 | 0.034 | 0.034 | 0.034 |

*Note:* $^{*}p<0.1$; $^{**}p<0.05$; $^{***}p<0.01$
Control for hospital fixed effects, number of beds, total budget, number of staff and proportion of admissions by age-sex group

#### 7.4 Total inpatient admissions

Table 34: Managerial quality and Total inpatient admissions 2012/13 – 2018/19

| | Total inpatient admissions | | | | | | | | | | | |
|---|---|---|---|---|---|---|---|---|---|---|---|---|
| | overall | 7a | 7b | 7c | 7d | 7e | 7f | 7g | 8a | 8b | 8c | 8d |
| | (1) | (2) | (3) | (4) | (5) | (6) | (7) | (8) | (9) | (10) | (11) | (12) |
| overall | −80.677 | | | | | | | | | | | |
| | (224.766) | | | | | | | | | | | |
| 7a | | −24.288 | | | | | | | | | | |
| | | (221.843) | | | | | | | | | | |
| 7b | | | −55.074 | | | | | | | | | |
| | | | (202.367) | | | | | | | | | |
| 7c | | | | −21.672 | | | | | | | | |
| | | | | (194.115) | | | | | | | | |
| 7d | | | | | −91.820 | | | | | | | |
| | | | | | (195.681) | | | | | | | |
| 7e | | | | | | −160.548 | | | | | | |
| | | | | | | (218.805) | | | | | | |
| 7f | | | | | | | 278.452 | | | | | |
| | | | | | | | (293.980) | | | | | |
| 7g | | | | | | | | 192.878 | | | | |
| | | | | | | | | (346.803) | | | | |
| 8a | | | | | | | | | −84.524 | | | |
| | | | | | | | | | (187.298) | | | |
| 8b | | | | | | | | | | −48.983 | | |
| | | | | | | | | | | (148.326) | | |
| 8c | | | | | | | | | | | −2.974 | |
| | | | | | | | | | | | (152.845) | |
| 8d | | | | | | | | | | | | 6.363 |
| | | | | | | | | | | | | (154.966) |
| Observations | 893 | 893 | 893 | 893 | 893 | 893 | 513 | 513 | 893 | 893 | 893 | 893 |
| $R^2$ | 0.605 | 0.605 | 0.605 | 0.605 | 0.605 | 0.605 | 0.438 | 0.437 | 0.605 | 0.605 | 0.605 | 0.605 |

*Note:* $^{*}$p<0.1; $^{**}$p<0.05; $^{***}$p<0.01

Control for hospital fixed effects, number of beds, total budget, number of staff and proportion of admissions by age-sex group

#### 7.5 Summary Hospital-level Mortality Indicator

Table 35: Managerial quality and Summary Hospital-level Mortality Indicator 2012/13 – 2018/19

| | Summary Hospital-level Mortality Indicator | | | | | | | | | | | |
|---|---|---|---|---|---|---|---|---|---|---|---|---|
| | overall | 7a | 7b | 7c | 7d | 7e | 7f | 7g | 8a | 8b | 8c | 8d |
| | (1) | (2) | (3) | (4) | (5) | (6) | (7) | (8) | (9) | (10) | (11) | (12) |
| overall | 0.0001<br>(0.001) | | | | | | | | | | | |
| 7a | | 0.001<br>(0.001) | | | | | | | | | | |
| 7b | | | 0.001<br>(0.001) | | | | | | | | | |
| 7c | | | | 0.0004<br>(0.001) | | | | | | | | |
| 7d | | | | | 0.0004<br>(0.001) | | | | | | | |
| 7e | | | | | | 0.001<br>(0.001) | | | | | | |
| 7f | | | | | | | 0.001<br>(0.002) | | | | | |
| 7g | | | | | | | | 0.001<br>(0.002) | | | | |
| 8a | | | | | | | | | 0.00003<br>(0.001) | | | |
| 8b | | | | | | | | | | −0.001<br>(0.001) | | |
| 8c | | | | | | | | | | | −0.001<br>(0.001) | |
| 8d | | | | | | | | | | | | −0.0005<br>(0.001) |
| Observations | 888 | 888 | 888 | 888 | 888 | 888 | 508 | 508 | 888 | 888 | 888 | 888 |
| $R^2$ | 0.034 | 0.035 | 0.035 | 0.034 | 0.034 | 0.035 | 0.050 | 0.050 | 0.034 | 0.035 | 0.034 | 0.034 |

*Note:* $^{*}p<0.1$; $^{**}p<0.05$; $^{***}p<0.01$

Control for hospital fixed effects, number of beds, total budget, number of staff and proportion of admissions by age-sex group
